# COVID-19 susceptibility and severity risks in a survey of over 500,000 individuals

**DOI:** 10.1101/2020.10.08.20209593

**Authors:** Spencer C. Knight, Shannon R. McCurdy, Brooke Rhead, Marie V. Coignet, Danny S. Park, Genevieve H.L. Roberts, Nathan D. Berkowitz, Miao Zhang, David Turissini, Karen Delgado, Milos Pavlovic, AncestryDNA Science Team, Asher K. Haug Baltzell, Harendra Guturu, Kristin A. Rand, Ahna R. Girshick, Eurie L. Hong, Catherine A. Ball

## Abstract

**Background:** The enormous toll of the COVID-19 pandemic has heightened the urgency of collecting and analyzing population-scale datasets in real time to monitor and better understand the evolving pandemic.

**Methods:** The AncestryDNA COVID-19 Study collected self-reported survey data on symptoms, outcomes, risk factors, and exposures for over 563,000 adult individuals in the U.S. in just under four months, including over 4,700 COVID-19 cases as measured by a self-reported positive test.

**Results:** We replicated previously reported associations between several risk factors and COVID-19 susceptibility and severity outcomes, and additionally found that differences in known exposures accounted for many of the susceptibility associations. A notable exception was elevated susceptibility for males even after adjusting for known exposures and age (adjusted odds ratio [aOR]=1.36, 95% confidence interval [CI] = (1.19, 1.55)). We also demonstrated that self-reported data can be used to build accurate risk models to predict individualized COVID-19 susceptibility (area under the curve [AUC]=0.84) and severity outcomes including hospitalization and critical illness (AUC=0.87 and 0.90, respectively). The risk models achieved robust discriminative performance across different age, sex, and genetic ancestry groups within the study.

**Conclusion:** The results highlight the value of self-reported epidemiological data to rapidly provide public health insights into the evolving COVID-19 pandemic.

**THUMBNAIL:** *What is already known on this subject:*

- The COVID-19 pandemic has exacted a historic toll on human lives, healthcare systems and global economies, with over 83 million cases and over 1.8 million deaths worldwide as of January 2021.
- COVID-19 risk factors for susceptibility and severity have been extensively investigated by clinical and public health researchers.
- Several groups have developed risk models to predict COVID-19 illness outcomes based on known risk factors.

*What this study adds:*

- We performed association analyses for COVID-19 susceptibility and severity in a large, at-home survey and replicated much of the previous clinical literature.
- Associations were further adjusted for known COVID-19 exposures, and we observed elevated positive test odds for males even after adjustment for these known exposures.
- We developed risk models and evaluated them across different age, sex, and genetic ancestry cohorts, and showed robust performance across all cohorts in a holdout dataset.
- Our results establish large-scale, self-reported surveys as a potential framework for investigating and monitoring rapidly evolving pandemics.

## INTRODUCTION

The COVID-19 pandemic has resulted in over 83 million COVID-19 cases and over 1.8 million deaths worldwide [1], including nearly 21 million cases and more than 350,000 deaths in the United States as of early January 2021 [2]. The growing impact of the pandemic intensifies the need for real-time understanding of COVID-19 susceptibility and severity risk factors, not only for public health experts, but also for individuals seeking to assess their own personalized risk. Prior research has indicated that differences in COVID-19 *susceptibility* are related to age [3], sex-dependent immune responses [4], and genetics [5, 6], while heightened *severity* of COVID-19 illness is associated with risk factors such as age [3,7–9], sex [4,10–12], genetic factors [13], and underlying health conditions [7,9,10,14,15]. Self-reported survey data, which can easily be collected in the home, afford the opportunity to dynamically monitor the continually evolving pandemic and allow for real-time estimation of individual-level COVID-19 risk [16–19]. Furthermore, self-reported surveys allow for collection of information about known exposures, of which few epidemiological COVID-19 studies have explicitly accounted for in association analyses to date [20].

In this paper, we aimed to replicate previous literature and to provide new insight into factors associated with susceptibility and severity of COVID-19 using a large survey cohort of 563,141 AncestryDNA customers who have consented to participate in the AncestryDNA COVID-19 Study [5]. We performed association tests of known or suspected COVID-19 risk factors with one susceptibility and two severity phenotypes and report unadjusted ORs and ORs adjusted for potential confounding factors. We additionally investigated associations of COVID-19 symptoms with susceptibility and severity.

We further demonstrate that this type of self-reported dataset can be used to build accurate predictive risk models for COVID-19 susceptibility and severity outcomes. For susceptibility, we designed two models and additionally applied two literature-based models [18] to predict COVID-19 cases among respondents reporting a test result. We also designed models to predict two different COVID-19 severity outcomes based on minimal information about demographics, health conditions, and symptoms: hospitalization due to COVID-19 infection and progression of an infection to a life-threatening critical case among those reporting a positive COVID-19 result [14]. To evaluate the potential for generalizability, we assessed performance of all of the risk models across different age, sex and genetic ancestry cohorts.

## METHODS

### Ethics approval

All data for this research project were from subjects who have provided informed consent to participate in AncestryDNA’s Human Diversity Project, as reviewed and approved by our external institutional review board, Advarra (formerly Quorum, IRB approval number: Pro00034516). Advarra operates under ethical principles underlying the involvement of human subjects in research, including the Declaration of Helsinki. All data were de-identified prior to use.

### Survey description

Survey responses were collected from AncestryDNA customers consented to research in the U.S. between April 22 and July 6, 2020. The survey consisted of 50+ questions about COVID-19 test results, 15 symptoms among those who tested positive or who tested negative and had flu-like symptoms, disease progression for positive testers, age, height, weight, known exposures to biological relatives, household members, patients or any other contacts with COVID-19, and 11 underlying health conditions (Supplementary Tables 1 and 2). Collection of self-reported COVID-19 outcomes from U.S. AncestryDNA customers who consented to research for the study, and the survey design are described in more detail in a genome-wide association study (GWAS) on a very similar AncestryDNA dataset [5]. Here, participants reporting a negative test result were also assessed for symptoms and clinical outcomes.

### Outcome definitions

The study assessed three outcomes: one for susceptibility and two for severity of COVID-19 infection. Cases for COVID-19 susceptibility were individuals who responded, “Yes, and was positive” to the question, “Have you been swab tested for COVID-19, commonly referred to as coronavirus?” Responders who answered, “Yes, and was negative” were used as controls for the susceptibility analysis.

The hospitalization outcome was defined among COVID-19-positive cases if a participant responded “Yes” to a binary question about experiencing symptoms due to COVID-19 illness and “Yes” to the hospitalization question (“Were you hospitalized due to these symptoms?). Controls were defined by a response of “No” to the symptoms question or a response of “No” to the hospitalization question in addition to reporting a self-reported positive COVID-19 test result [5].

Critical cases of COVID-19 were defined via a response of “Yes” to one or more questions about ICU admittance or, alternatively, self-reported septic shock, organ failure, or respiratory failure resulting from a COVID-19 infection [14]. Controls were defined by a response of “No” across all of these questions in addition to self-reporting a positive COVID-19 test result.

### Genetic sex and ancestry definitions

All individuals were genotyped, using previously described general genotyping and quality control procedures [21]. Both sex and genetic ancestry were defined for individuals based on their genotypes. Genetic ancestry was estimated using a proprietary algorithm to estimate continental admixture proportions [22]. All participants were assigned to one of four broad genetic ancestry groups: European ancestry, admixed African-European ancestry, admixed Amerindian ancestry, or other ancestry combinations.

### Data preparation for analysis

Multiple-choice categorical questions were one-hot (“dummy”) encoded as binary risk factors. We considered several risk factors and outcomes questions in our association analyses and risk modeling efforts, some of which are summarized in Supplementary Tables 1 and 2. Based on the dependency structure of the survey, we made the following inferences:

- Participants reporting “No” to a binary question about symptoms arising from COVID-19 infection were designated as negatives for dependent questions about individual symptoms, hospitalization due to symptoms, and ICU admittance due to symptoms.
- Participants reporting “No” to a binary question about hospitalization were assigned to a hospital duration of 0 days and designated as negative for ICU admittance due to symptoms.

For association analyses, responses to a question about individual symptoms (“Between the beginning of February 2020 and now, have you had any of the following symptoms?”) were converted to a binary variable based on the following mapping: 0 = None, Very Mild, Mild; 1 = Moderate, Severe, Very Severe.

### Association analysis

Analyses were performed either with the *statsmodels* package in Python3 or in base R with the *glm* function. For each susceptibility and severity outcome and risk factor of interest, a simple logistic regression (LR) model was fit using unpenalized maximum likelihood (Supplementary Tables 6-14) [23]. Multiple logistic regression was used to adjust the ORs for known COVID-19 exposures and potentially confounding risk factors. The adjusted model included age, sex, and four known exposures (Y/N if any) for susceptibility outcomes; and age, sex, obesity (binarized if BMI >= 30), and health conditions (binarized if any) for severity outcomes (Figures 1 and 2). Individual adjustment variables were omitted when analyzing associations for risk factors within equivalent categories (e.g., age was not included in adjusted models for age bin risk factors).

**Figure 1.**
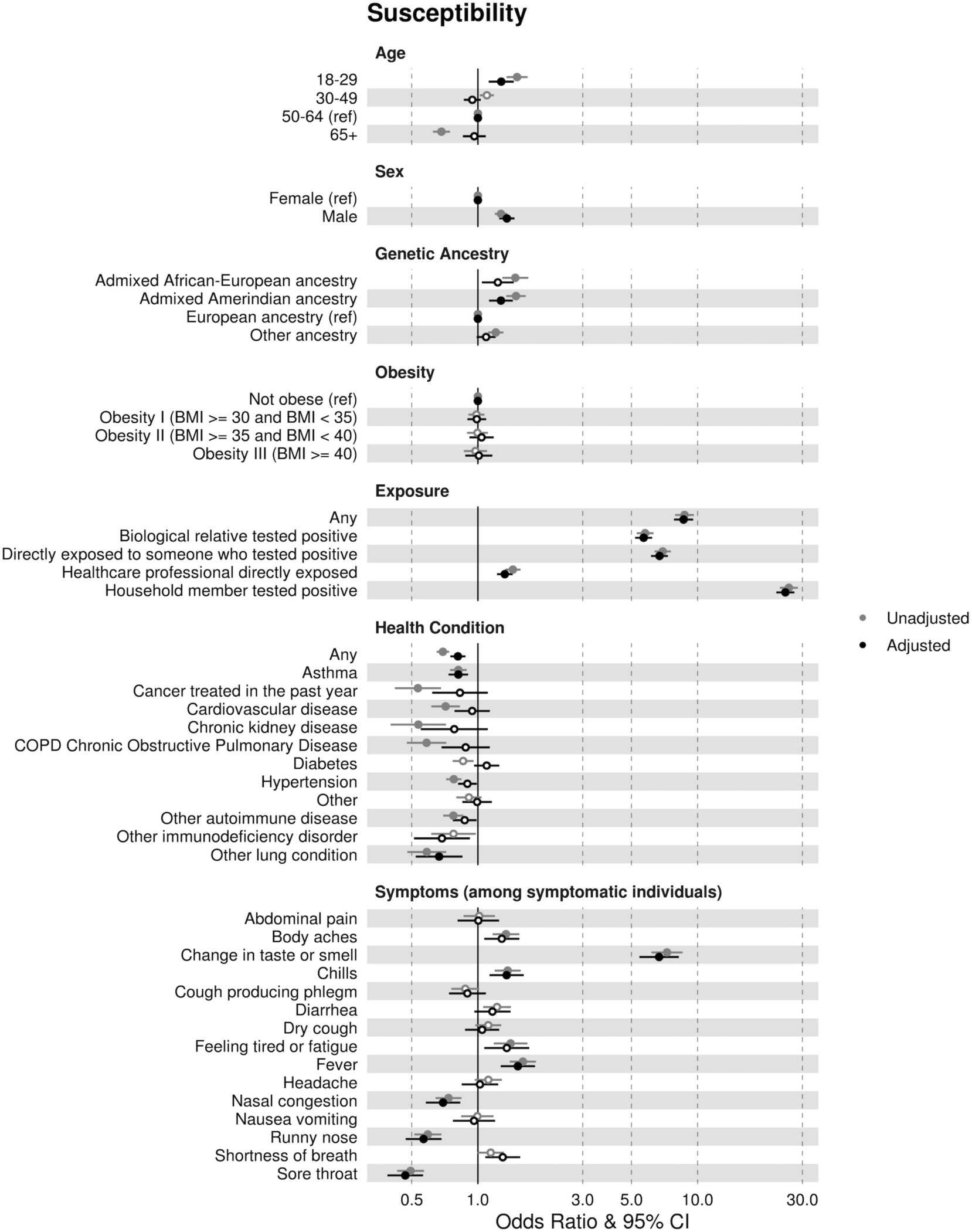
Susceptibility odds ratios (ORs) and 95% confidence intervals (CIs) estimated from simple (“Unadjusted models,” grey) and multiple (“Adjusted models,” black) logistic regression with adjustment for other risk factors. Open circles indicate not significant (p-value > 0.05) after accounting for multiple hypothesis tests using Bonferroni correction. Age, sex, genetic ancestry, and obesity ORs were estimated in relation to the reference variables indicated. Exposure, health, and symptom ORs were each estimated separately as binary variables. Symptom ORs were estimated as binary variables among symptomatic testers only (Methods). Risk factor adjustments for susceptibility include: sex, age, and at least one known COVID-19 exposure. Where applicable, individual adjustment variables were omitted to avoid duplicate adjustment (Methods).

**Figure 2.**
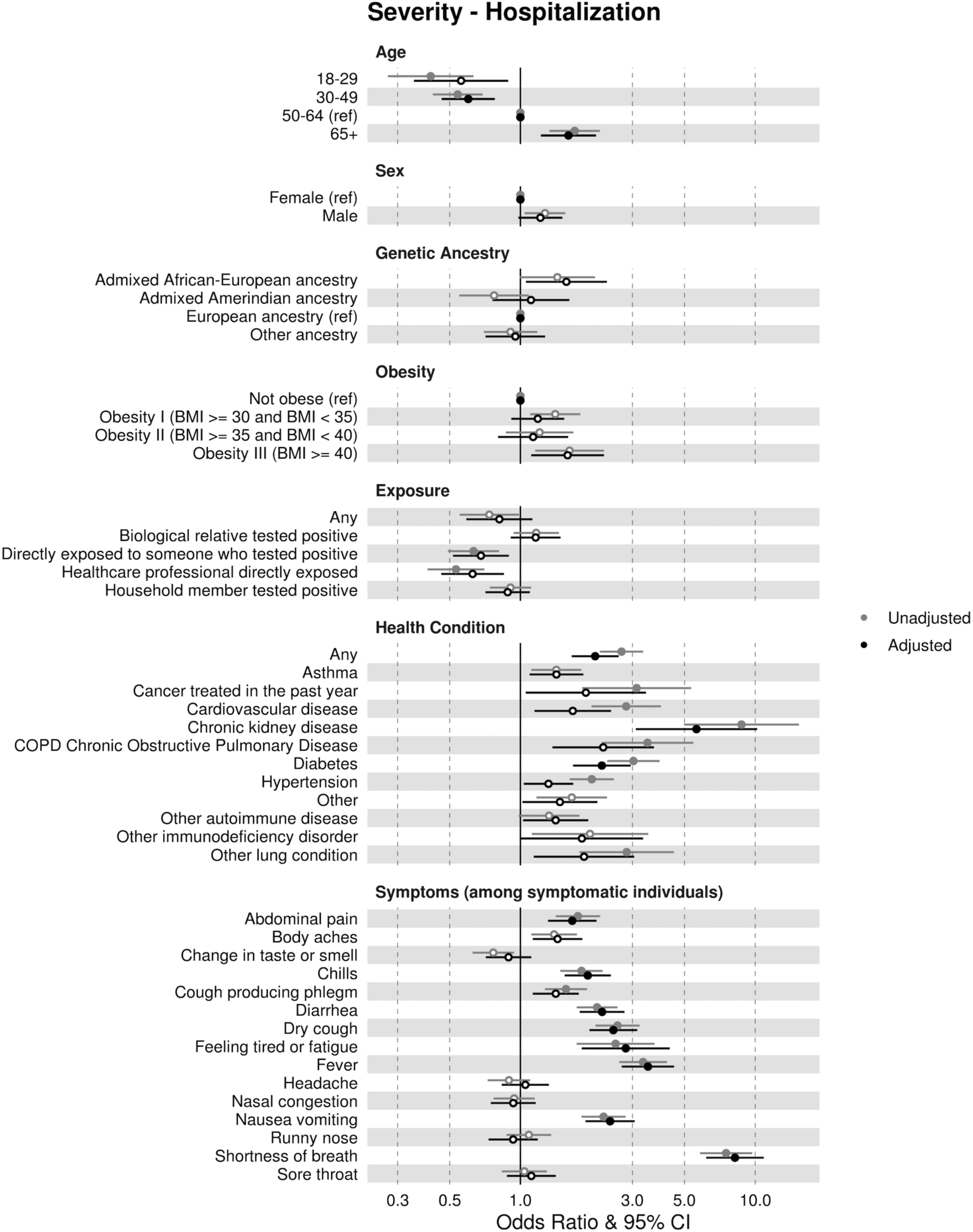
Severity (hospitalization) odds ratios (ORs) and 95% confidence intervals (CIs) estimated from simple (“Unadjusted models,” grey) and multiple (“Adjusted models,” black) logistic regression with adjustment for other risk factors. Open circles indicate not significant (p-value > 0.05) after accounting for multiple hypothesis tests using Bonferroni correction. Age, sex, genetic ancestry, and obesity ORs were estimated in relation to the reference variables indicated. Exposure, health, and symptom ORs were each estimated separately as binary variables. Symptom ORs were estimated as binary variables among symptomatic testers only (Methods). Risk factor adjustments for severity include: sex, age, obesity (Y/N), and underlying health conditions (Y/N if any). Where applicable, individual adjustment variables were omitted to avoid duplicate adjustment (Methods). See Supplementary Figure 1 for critical case severity ORs.

For each risk factor, 95% confidence intervals (CIs) for the log odds ratio were estimated under the normal approximation. The significance threshold was Bonferroni-corrected for the 42 different risk factors examined (adjusted threshold of 0.05/42=0.0012) [23].

### Risk factor selection and risk model training

Prior to model training, the data were split with a fixed random seed into training and holdout datasets. We chose risk factors based on a minimal subset of nominally significant ORs within our training data as well as literature guidance [3,4,7,9,11,12,14,15]. For the susceptibility models without symptoms, we included a subset of exposure-related questions, based on the training OR analyses, as well as two demographic variables (age and sex, Supplementary Tables 15-17). For susceptibility models with symptoms, we additionally included the five symptoms most differentiated between symptomatic negative and positive testers from our training ORs. For the severity models, we included pre-existing conditions, based on the training OR analyses, predictive symptoms within our training dataset, morbid obesity (BMI >= 40), age, and sex (Supplementary Table 18).

Once final risk factors were selected, we trained LR models with 5-fold cross-validated grid search on the training dataset to select an optimal lasso regularization parameter lambda [23]. For the grid search, we scanned 8 different values for lambda, equally partitioned geometrically across a 4-log space. We then re-trained on the entire training dataset with the optimal lambda and evaluated the final model on the holdout dataset.

### Model thresholding

Phenotypes were predicted from the output of trained models based on a 50% probability threshold (i.e., logistic model output > 0.5). Sensitivity and specificity were then calculated based on the true vs. predicted binary outcomes.

### Estimation of performance error

To estimate error in model performances, we bootstrapped our holdout dataset 1,000 times to generate a sampling distribution for each evaluation metric. We estimated the mean and 95% CIs for each metric based on the mean and standard deviation of this sampling distribution [23].

## RESULTS

### Survey response and study population

A total of 563,141 responses were collected, with 4,726 individuals reporting a COVID-19 positive test result, 28,872 a negative test result, 71,761 no COVID-19 test but flu-like symptoms, and 454,542 no COVID-19 and no flu-like symptoms. 3,240 reported pending test results and were excluded from further analyses. The survey completion rate was approximately 95%. In general, the COVID-19 positive test rate and self-reported clinical outcomes were consistent with those reported by the U.S. Centers for Disease Control and Prevention (CDC) over a similar period (Supplementary Note 1) [24]. The majority of participants were female (67.5%) and of European ancestry (75.4%), with some individuals of Admixed Amerindian (6.5%) or Admixed African-European (3.0%) ancestries. The median age of the entire cohort was 56, and the median age of those reporting a positive test result was 49 (Table 1 and Supplementary Tables 1-5).

**Table 1.** Study population demographic information.

|  | COVID-19<br>Nasopharyngeal<br>Swab Test<br>Positive<br>*n* = 4,726 | COVID-19<br>Nasopharyngeal<br>Swab Test<br>Negative<br>*n* = 28,872 | Full AncestryDNA<br>COVID-19 Cohort<br>*n* = 563,141 |
| --- | --- | --- | --- |
| **Age** |  |  |  |
| Median, mean (sd) | 49, 49.49 (15.43) | 53, 52.68 (15.5) | 56, 53.90 (16.13) |
| Bins (Counts, %) |  |  |  |
| 18-30 | 600 (0.13) | 2496 (0.09) | 52580 (0.09) |
| 31-40 | 941 (0.2) | 5001 (0.17) | 87261 (0.15) |
| 41-50 | 926 (0.2) | 5333 (0.18) | 90473 (0.16) |
| 51-60 | 1057 (0.22) | 6009 (0.21) | 108062 (0.19) |
| 61-70 | 735 (0.16) | 5961 (0.21) | 128200 (0.23) |
| 71-90 | 467 (0.10) | 4072 (0.14) | 96565 (0.17) |
| **Genetic Sex (counts, %)** |  |  |  |
| Female | 3013 (0.64) | 19945 (0.69) | 380349 (0.67) |
| Male | 1706 (0.36) | 8867 (0.31) | 183153 (0.32) |
| **Genetic Ancestry Continental Groupings** |  |  |  |
| Admixed African-European ancestry | 275 (0.06) | 1244 (0.04) | 17019 (0.03) |
| Admixed Amerindian ancestry | 520 (0.11) | 2336 (0.08) | 36865 (0.07) |
| European ancestry | 3026 (0.64) | 20269 (0.7) | 424328 (0.75) |
| Other ancestry | 905 (0.19) | 5023 (0.17) | 84929 (0.15) |
| **Pre-existing health conditions (any)** | 1911 (0.46) | 15261 (0.55) | 255788 (0.47) |
| **BMI (med, mean, sd)** | 28.59, 29.83 (7.04) | 28.67, 29.92 (7.05) | 28.29, 29.53 (6.87) |
| **Underweight (count, %)** | <100 | 204 | 4407 |
| **Healthy (count, %)** | 971 | 6442 | 139565 |
| **Overweight (count, %)** | 1225 | 8503 | 172941 |
| **Obese (count, %)** | 1629 | 11264 | 203325 |
| **Flu** |  |  |  |
| Shot (yes) | 2656 (0.63) | 19856 (0.71) | 356144 (0.65) |
| Test Dx (yes) | 198 (0.05) | 1180 (0.04) | 11726 (0.02) |
| **Symptoms (tested pos.)** |  |  |  |
| General, yes (count, % of pos.) | 3862 (0.87) |  | 9237 (0.47) |
| **Hospitalization (count, % of positives)** | 453 (0.11) |  | 1397 (0.02) |
| Duration, days (med, mean, sd) | 5, 7.62 (9.08) |  | 3, 4.78 (7.22) |

### Susceptibility associations: replicated and novel

We replicated many previously reported literature associations for susceptibility. The strongest associations for a positive COVID-19 test result were known COVID-19 exposures, either through a household case (OR=26.03; 95% confidence interval [CI]=(22.26, 30.43)), biological relative (OR=5.77; 95% CI=(4.99, 6.68)), or other source of “direct” exposure (OR=6.94; 95% CI=(6.02, 7.99)) (Figure 1, Supplementary Table 6). In general, adjusting for known exposures, age, and sex resulted in attenuation of the ORs, with many associations becoming insignificant after adjustment (Figure 1, Supplementary Table 7).

One novel result was that the OR for males was not attenuated after adjustment, and males remained at elevated odds after adjusting for known exposures and age (aOR=1.36; 95% CI=(1.19, 1.55); Figure 1, Supplementary Table 7). We also note that males and females reported comparable exposure burden, with males slightly more likely to report a household case of COVID-19 but less likely to report a case of COVID-19 among biological relatives (Supplementary Tables 9 and 10).

Consistent with previous reports [25–29], younger individuals (ages 18-29; OR=1.51; 95% CI=(1.26, 1.81)) were significantly more likely to test positive compared to older individuals (ages 50-64, the largest age group in this cohort), and individuals of admixed African-European (OR=1.48; 95% CI=(1.18, 1.85)) or admixed Amerindian ancestry (OR=1.49; 95% CI=(1.26, 1.77)) were more likely to test positive compared to those of European ancestry (Supplementary Table 6). Individuals in all three of these groups reported higher levels of COVID-19 cases within the household, cases among biological relatives, and/or other known “direct” COVID-19 exposures (Supplementary Tables 8-10). Adjusting for age (ancestry groups only), sex, and known exposures attenuated the OR for all of these groups (younger aOR=1.28; 95% CI=(1.03, 1.59), African-European aOR=1.23; 95% CI=(0.94, 1.62), and Amerindian aOR=1.27; 95% CI=(1.04, 1.57); Figure 1, Supplementary Table 7).

Individuals reporting pre-existing medical conditions (e.g., cancer, cardiovascular disease, chronic kidney disease [CKD], diabetes, hypertension) were less likely to test positive for COVID-19 (Figure 1, Supplementary Table 6). We observed significantly decreased odds of a known “direct” exposure to COVID-19, as well as significantly decreased odds of a household case of COVID-19, among such individuals relative to those without any health conditions (OR=0.71; 95% CI=(0.65, 0.78) and OR=0.74; 95% CI=(0.65, 0.84), respectively; Supplementary Tables 8 and 9).

### Replicated associations for COVID-19 severity

Consistent with previous reports [7,9,12,14,15], we observed positive associations between certain health conditions and COVID-19 severity outcomes; many of these associations remained significant after adjustment for age, sex, and obesity (BMI >= 30) (Figure 2, Supplementary Tables 11-14). COVID-19 cases reporting at least one underlying health condition were significantly more likely to progress to a critical case (OR=2.85; 95% CI=(1.78, 4.57); Figure 2, Supplementary Figure 1, Supplementary Table 13). Specific underlying health conditions that were associated with hospitalization and/or critical case progression included CKD, chronic obstructive pulmonary disease (COPD), diabetes, cardiovascular disease, and hypertension (Figure 2, Supplementary Figure 1, Tables S12 and S14). Among individuals testing positive for COVID-19, the oldest (≥65 years) were significantly more likely to be hospitalized compared to those aged 50-64 (OR=1.70; 95% CI=(1.13, 2.56); Figure 2, Supplementary Table 11). Individuals of admixed African-European ancestry who tested positive were significantly more likely to report progression to a critical case, compared to those with European ancestry (OR=2.07; 95% CI=(1.03, 4.17); Supplementary Figure 1, Supplementary Table 13). Among COVID-19 cases, males were significantly more likely than females to report progression to a critical case (OR=1.54, 95%; CI=(1.00, 2.37); Supplementary Figure 1, Supplementary Table 13); these findings are consistent with CDC reports of increased ICU admittance rates in males (3% vs. 2%) [24].

### Differential symptomology between susceptibility and severity

We compared associations between susceptibility and severity to provide a more nuanced view of symptoms associated with a positive test result versus those associated with severe illness progression (Figure 3) [17,18,30]. Among symptomatic people reporting a COVID-19 test result, those reporting change in taste or smell (OR=7.26; 95% CI=(5.54, 9.50)), fever (OR=1.60; 95% CI=(1.28, 2.01)), or feeling tired or fatigue (OR=1.41; 95% CI=(1.05, 1.89)) were more likely to test positive (Figure 3, Supplementary Table 6). Those reporting runny nose (OR=0.59; 95% CI=(0.47, 0.75)) or sore throat (OR=0.49; 95% CI=(0.39, 0.62)) were more likely to test negative, consistent with previous reports that these symptoms are more indicative of influenza or the common cold (Figure 3, Supplementary Table 6) [17,18,30]. Change in taste or smell, a hallmark symptom of COVID-19 infection, was not associated with hospitalization (OR=0.77, 95% CI=(0.55, 1.07); Figure 3, Supplementary Table 11). By contrast, dyspnea (shortness of breath) was strongly associated with hospitalization and critical case progression (OR=7.52; 95% CI=(4.92, 11.49) and OR=11.55; 95% CI=(5.91, 22.59), respectively) [31], but was not associated with a positive test result (OR=1.14; 95% CI=(0.91, 1.44); Figure 3, Supplementary Tables 6, 11, and 13).

**Figure 3.**
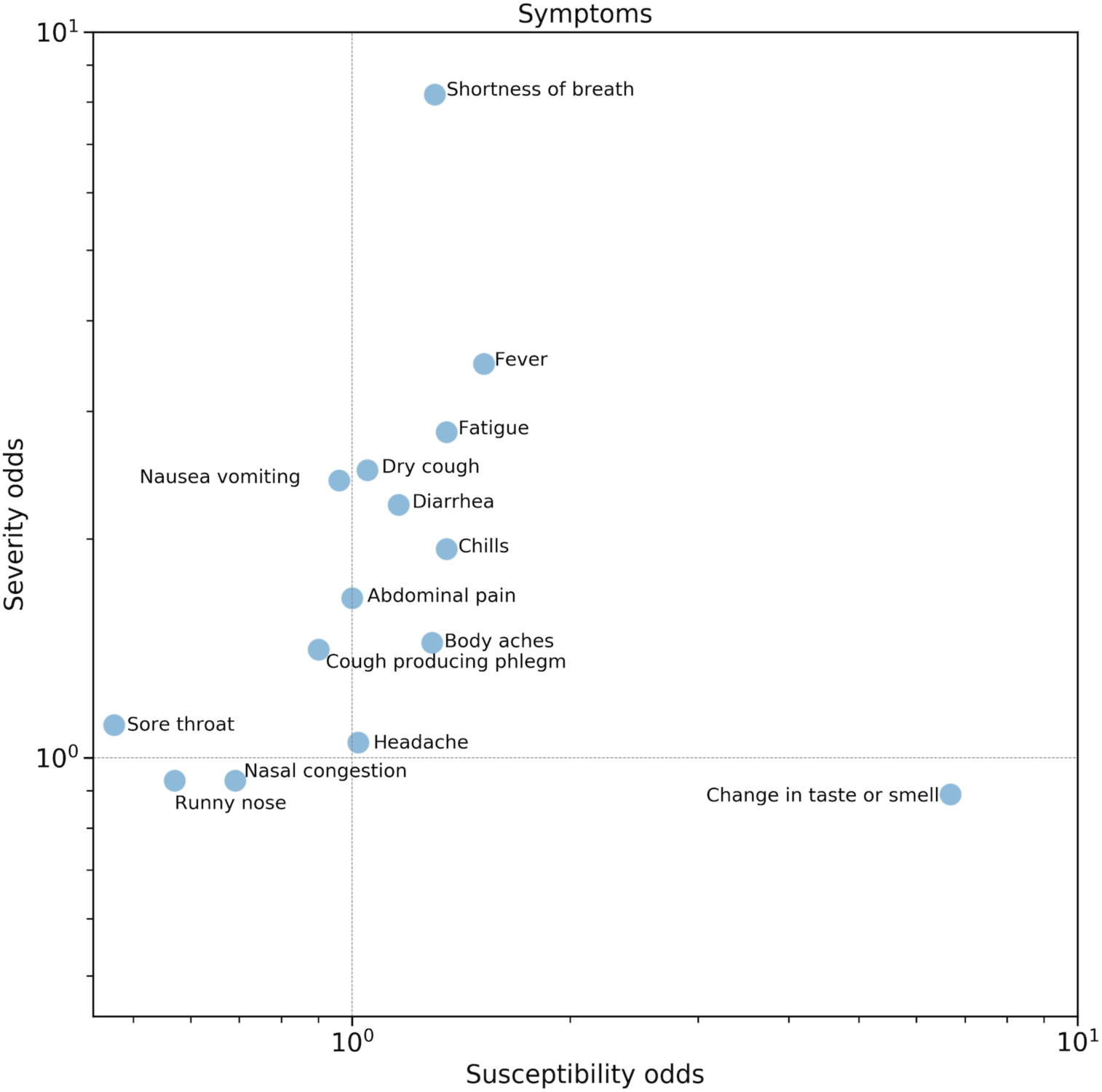
Comparison of susceptibility adjusted ORs (horizontal axis) and severity adjusted ORs (vertical axis) for symptoms in Figures 1 and 2. Severity aORs are for hospitalization. Note that aORs for susceptibility and severity are adjusted differently according to descriptions in Figure 1 and 2 captions. Adjusted ORs are plotted on a log scale for visibility. Shortness of breath is the strongest indicator of increased severity, while change in taste or smell is the strongest indicator for testing positive for COVID-19 among symptomatic individuals (Methods). Refer to Supplementary Figure 2 for demographic, health condition, and exposure aORs.

### Predictive risk models

We further developed risk models that predict an individual’s COVID-19 risk (positive test result or severity, Methods) [7,16–18,32,33]. The susceptibility models were designed to predict a COVID-19 result (positive or negative) from risk factors among testers. We compared four models: our model based on demographics and exposures only (“Dem + Exp”); our model based on demographics, exposures, and symptoms (“Dem + Exp + Symp”); and for benchmarking purposes, a replication of a previously published model called “How We Feel” based on nearly identical self-reported symptoms (“HWF Symp”), and one which also included self-reported exposures (“HWF Exp + Symp”) (Supplementary Note 2, Supplementary Table 18) [18].

All four susceptibility models performed robustly; the three models that included one or more symptoms outperformed the model without symptoms (Dem + Exp), underscoring the value of self-reported symptoms for discriminating between cases and controls (Figure 4, see Supplementary Tables 19-22 for detailed model performance data). The model with demographics, exposures, and symptoms (Dem + Exp + Symp) achieved the highest overall performance with an AUC of 0.94 ± 0.02, a sensitivity of 85%, and a specificity of 91% (Supplementary Note 3). Each of the models performed comparably across different age, sex, and genetic ancestry cohorts (Figure 4, Supplementary Tables 19-22). We observed no significant overfitting in any of the models as evidenced by comparable train-test performances (Supplementary Table 23).

**Figure 4.**
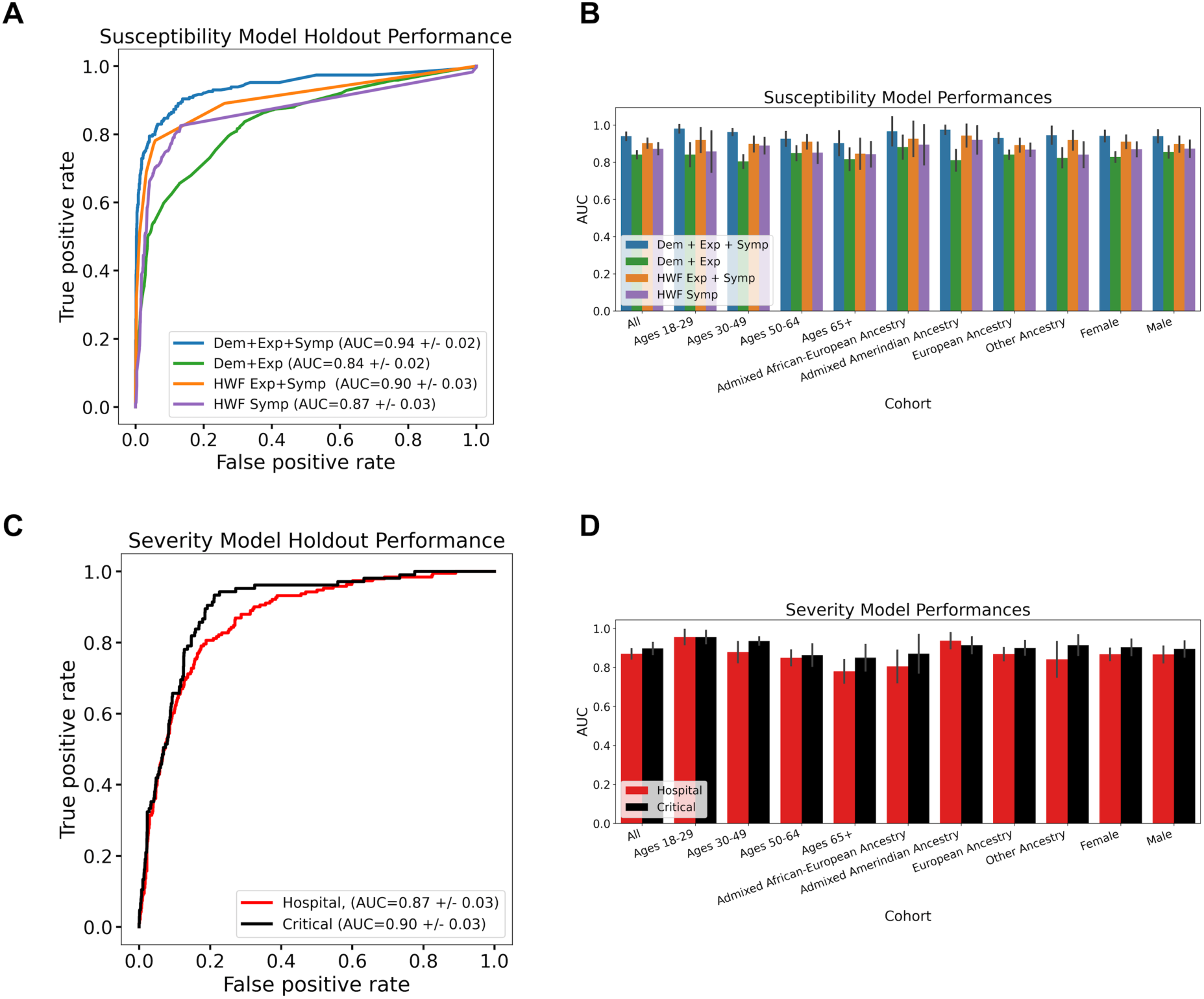
Performance of risk models on independent holdout data. A) Receiver operating characteristic (ROC) curves for susceptibility models to predict COVID-19 cases among testers reporting a result (positive or negative). B) Area under the curve (AUC) for the four susceptibility models in A), stratified by cohort. “All” represents everyone in A). C) ROC curves for severity models to predict either hospitalization (red) or critical illness progression (black) among COVID-19 cases. D) Area under the curve (AUC) for the two severity models in B), stratified by cohort. “All” represents everyone in C). Refer to Methods as well as Supplementary Figure 3 and Supplementary Tables 18-25 for additional model performance data and model risk factor information.

We trained two severity models, designed to predict either hospitalization or progression to critical illness among COVID-19 cases. We included a number of risk factors and symptoms most associated with severe COVID-19 outcomes from the literature and/or our training dataset (Figure 3, Supplementary Tables 16 and 17); these included age [7–9,14], sex [4,7,11,12,14], morbid obesity (BMI >= 40) [7, 34], and health conditions [7,9,12,14,15], as well as symptoms including shortness of breath [31], fever, feeling tired or fatigue, dry cough, and diarrhea. Both models performed robustly on an independent holdout dataset (AUCs of 0.87 +/− 0.03 and 0.90 +/− 0.03 for the hospitalization and critical models, respectively, Figure 4). The severity models performed comparably when stratifying by age, sex, and genetic ancestry (Figure 4, Supplementary Tables 24 and 25), and there was no significant overfitting bias as evidenced by comparable train-test performances (Supplementary Table 23).

## DISCUSSION

The AncestryDNA COVID-19 Study provides a highly complete, self-reported dataset that contains information about a plethora of risk factors in the context of COVID-19 susceptibility and severity outcomes. The self-report framework provides fast, low-cost, population-scale data that are particularly valuable in a pandemic, where knowledge is both limited and evolving rapidly based on changing circumstances. Additionally, the broad collection mechanism enables data-gathering from many more participants than typically seen in a medical setting, including those with mild or no symptoms, and participants can safely provide data from their homes.

The study highlights exposure burden as the primary risk factor for COVID-19 susceptibility, and the importance of accounting for known exposures when assessing differences in susceptibility to COVID-19. Few studies have measured and explicitly adjusted for known COVID-19 exposures at this scale [20]. Importantly, we found elevated susceptibility risk in males after adjusting for age and known exposures, and unlike most of the risk factors we evaluated, the adjusted odds were not attenuated compared to the unadjusted odds. This finding is distinct from previous findings on elevated severity risk in males [4,7,11]. This result could be due to differences between men and women in behaviors, unknown exposures, biology, genetics [4–6], or other risk factors not measured within this dataset and should be investigated in future studies.

Another major contribution of this study is the use of self-reported data for the development of novel risk models for predicting an individual’s COVID-19 susceptibility and severity risk. The risk models presented here perform comparably or better than similar and more complex models reported previously [16–18,32,33]. Although some previously reported risk models have been assessed in different age or sex cohorts [16, 17], we are not aware of any that have been assessed across genetic ancestry cohorts [7,16–18,32,33], highlighting the potential utility and generalizability of the models to broader populations [17,18,30].

### Limitations

We note that there are some inherent limitations of self-reported data for studying COVID-19 risk factors. The most severe cases, especially those resulting in mortality, were not sampled. As a result, many of the risk factor effect estimates may be underestimated. Additionally, the AncestryDNA cohort is self-selected, slightly older, more European, and more female than the broader U.S. population. Another potential issue is that those who reported a negative test may have underestimated their exposures and symptoms relative to those who tested positive, leading to upwardly biased exposure effect estimates. Finally, misclassification of COVID-19 positive status is likely given the uneven availability of tests over the time period surveyed, potentially leading to susceptibility effect estimates that are biased toward the null. However, the fact that most of the associations observed in this study were similar to those previously reported in the literature and the fact that risk model performance remained high when data were stratified by age, sex, and genetic ancestry lends confidence to our findings in spite of limitations.

## CONCLUSION

The COVID-19 pandemic has exacted a historic toll on healthcare systems and global economies and continues to evolve based on changes in human behavior, public health guidelines, and societal factors. This study demonstrates the power of a self-reported data in a large cohort to rapidly elucidate more details about COVID-19 risk factors and help point the way to minimizing disease burden.

## Data Availability

A dataset (EGAC00001001762) is available to qualified scientists through the European Genome-phenome Archive (EGA). The EGA dataset includes the risk factors and outcomes studied here. The EGA dataset is de-identified and comprises ~15,000 individuals who tested for COVID-19, including more than 3,000 individuals who tested positive, many of whom are in this study. The EGA cohort is sufficient to nominally replicate the vast majority of susceptibility and severity associations from this study.

https://ega-archive.org/dacs/EGAC00001001762

## Abbreviations

aOR: adjusted odds ratio
AUC: area under the curve
BMI: body mass index
CI: confidence interval
CDC: U.S. Centers for Disease Control
CKD: chronic kidney disease
COPD: chronic obstructive pulmonary disease
EGA: European Genome-phenome Archive
GWAS: genome-wide association studies
HWF: “How We Feel” study
ICU: intensive care unit
LR: logistic regression
OR: odds ratio
ROC: receiver operating characteristic

## Acknowledgements

We thank our AncestryDNA customers who made this study possible by contributing information about their experience with COVID-19 through our survey. Without them, this work would not be possible. We would like to thank Zach Bass, Robert Dowling, Disha Akarte, Swapnil Sneham, Sean Enright and the entire Cyborg team for their tireless work in the release and continued support of the COVID-19 survey.

## Contributors

SCK, SRM, BR contributed equally to the manuscript and wrote the first draft of the paper. ARG provided direct project guidance and lead the COVID-19 research teams. SCK, SRM, BR performed the association analyses. SCK developed and assessed the risk models. MVC and KAR designed the COVID-19 survey questionnaire. SCK and DP supported the dataset creation. GHLR supported the phenotype definitions. MVC and NDB built the demographic tables. SCK, SRM, BR, MVC, GHLR, ARG, and KAR helped with additional analyses and interpretation. MZ, DP, DT, KD, MP, HG, AKHB helped with the EGA dataset. The AncestryDNA Science Team contributed to additional work, allowing for the completion of the COVID-19 research and manuscript. KAR, ELH, and CAB provided additional project guidance. All authors contributed to the final manuscript.

## AncestryDNA Science Team

Yambazi Banda, Ke Bi, Robert Burton, Marjan Champine, Ross Curtis, Abby Drokhlyansky, Ashley Elrick, Cat Foo, Michael Gaddis, Jialiang Gu, Shannon Hateley, Heather Harris, Shea King, Christine Maldonado, Evan McCartney-Melstad, Alexandra McFarland, Patty Miller, Luong Nguyen, Keith Noto, Jingwen Pei, Jenna Petersen, Scott Pew, Chodon Sass, Josh Schraiber, Alisa Sedghifar, Andrey Smelter, Sarah South, Barry Starr, Cecily Vaughn, Yong Wang

## Patient consent for publication

Not required.

## Data availability statement

A dataset (EGAC00001001762) is available to qualified scientists through the European Genome-phenome Archive (EGA). The EGA dataset includes the risk factors and outcomes studied here. The EGA dataset is de-identified and comprises ∼15,000 individuals who tested for COVID-19, including more than 3,000 individuals who tested positive, many of whom are in this study. The EGA cohort is sufficient to nominally replicate the vast majority of susceptibility and severity associations from this study. Risk models trained within the EGA cohort achieve comparable discriminative performance to the models presented here when evaluated in an independent holdout dataset (Supplementary Figure 5).

## SUPPORTING INFORMATION

### SUPPLEMENTARY NOTES

**Supplementary Note 1. Comparison of AncestryDNA data to CDC data.** In general, the COVID-19 testing results are consistent with those reported by the U.S. Centers for Disease Control and Prevention (CDC) over a similar period [24]. For example, the CDC reported a 12% cumulative overall positive test rate from 1 March to 30 May 2020, while 14.1% of AncestryDNA survey participants reported testing positive (the denominator includes tests with “results pending”). For hospitalization, the CDC reported that 14% of individuals testing positive for COVID-19 were hospitalized, and 2% were admitted to an intensive care unit (ICU), between 22 January and 30 May 2020. Among the AncestryDNA survey respondents, 11% of those reporting a positive COVID-19 test were hospitalized, and 4.9% were admitted to ICU between April 22 and July 6, 2020.

**Supplementary Note 2. Risk factors for predictive susceptibility models.** The HWF Exp + Symp model includes three risk factors, including two exposures and one symptom (change in taste or smell, Supplementary Table 18). The HWF Symp model includes seven symptoms most commonly associated with COVID-19 according to the CDC (Supplementary Table 18) [18]. For the Dem + Exp and Dem + Exp + Symp models, we incorporated responses to several exposure questions that were primary risk factors in the OR analysis of the training set (Methods and Supplementary Table 15) [18]. We also considered age and sex, given their nominal association with COVID-19 in our ORs in the training dataset, as well as reports about these variables as risk factors (Supplementary Table 15) [3,4,29]. For Dem + Exp + Symp model, we selected the most differentiated symptoms between cases and controls amongst symptomatic COVID-19 testers within the training dataset, including fever, change in taste or smell, feeling tired/fatigued, runny nose, and sore throat [17,18,30].

**Supplementary Note 3. Comparison of AncestryDNA and HWF cohorts and performances.** The HWF models performed better when trained and evaluated on the AncestryDNA dataset as compared with the HWF dataset, particularly for the symptoms-only model (HWF Symp: AncestryDNA-trained AUC=0.87 and HWF-trained AUC=0.76 and HWF Exp + Symp: AncestryDNA-trained AUC=0.90 and HWF-trained AUC=0.87; Figure 4) [18]. This result could be due to differences in survey flow design, demographics, or the relative prevalences of self-reported symptoms between the two datasets. We re-trained and evaluated our models on a subset of symptomatic COVID-19 testers (positives and negatives) reporting at least one symptom of moderate or greater intensity. The performances of all models that included symptoms were slightly attenuated in this cohort, with AUCs for the HWF models approaching previous reports (HWF Symp: AncestryDNA-trained AUC=0.80 and HWF Exp + Symp: AncestryDNA-trained AUC=0.87); the model without symptoms performed the same (Supplementary Figure 4). This suggests that predicting COVID-19 cases on the basis of symptoms alone is more challenging when symptoms are less differentiated between positive and negative testers.

### SUPPLEMENTARY TABLES

**Supplementary Table 1.** Selected subset of survey questions considered in the context of COVID-19 severity outcomes analysis and modeling efforts.

| Risk factor description | Question | Possible responses |
| --- | --- | --- |
| **COVID-19 test result** | Have you been swab tested for COVID-19, commonly referred to as coronavirus? | Yes, and was positive; Yes, and was negative; Yes, and my results are pending; No, but I have had flu-like symptoms with a fever at some point since the beginning of February 2020; No, and I have not had flu-like symptoms at some point since the beginning of February 2020 |
| **Case amongst direct biological relatives (multi-choice)** | Have any of your biological relatives tested positive for COVID-19? Select all that apply. | Grandparent(s); Parent(s); Child(ren); Sibling(s); Other; None |
| **Healthcare worker routinely exposed to COVID-19** | Are you a healthcare professional working directly with or in close physical proximity to patients who have tested positive for COVID-19? | Yes; No; Not sure |
| **Household case of COVID-19** | Has someone in your household tested positive for COVID-19? | Yes; No; Not sure |
| **Number of household cases of COVID-19** | How many people in your household have tested positive for COVID-19? | Numeric |
| **Known direct COVID-19 exposure** | Have you been directly exposed to someone who has tested positive for COVID-19? | Yes; No; Not sure |
| **Health condition(s)** | Do you currently have any of the following health conditions? Select all that apply. | Asthma, COPD (Chronic Obstructive Pulmonary Disease); Other lung condition; Cancer (treated in the past year); Cardiovascular disease; Chronic kidney disease; Diabetes; Hypertension; Organ failure requiring a transplant (in the last year); Blood disorder requiring hematopoietic stem cell / bone marrow transplant; Other autoimmune disease; Other immunodeficiency disorder; Other; None; Not sure |

**Supplementary Table 2.** Selected risk factors considered in susceptibility or severity outcomes analysis and modeling efforts.

| Risk factor description | Question | Possible responses |
| --- | --- | --- |
| **COVID-19 test result** | Have you been swab tested for COVID-19, commonly referred to as coronavirus? | Yes, and was positive; Yes, and was negative; Yes, and my results are pending; No, but I have had flu-like symptoms with a fever at some point since the beginning of February 2020; No, and I have not had flu-like symptoms at some point since the beginning of February 2020 |
| **Symptoms (binary)** | Did you experience symptoms as a result of your condition? | Yes; No; Not sure |
| **Symptoms (multi-choice)** | Between the beginning of February 2020 and now, have you had any of the following symptoms? | Fever; Shortness of breath; Dry cough; Nasal congestion; Runny nose; Sore throat; Feeling tired or fatigue; Chills; Body aches; Headache; Cough producing phlegm; Abdominal pain; Nausea or vomiting; Diarrhea; Change in taste or smell |
| **Symptom duration (multi-choice)** | How long did these symptoms last? | Less than 1 day; 1-3 days; 4-7 days; 8-14 days; 2 weeks to a month; 1-3 months; More than 3 months |
| **Hospitalization** | Were you hospitalized due to these symptoms? | Yes; No; Not sure |
| **Hospital duration** | How long were you hospitalized? | Numeric |
| **ICU with ventilator** | Were you hospitalized in the Intensive Care Unit (ICU) with a ventilator? | Yes; No; Not sure |
| **ICU with oxygen** | Were you hospitalized in the Intensive Care Unit (ICU) with oxygen? | Yes; No; Not sure |
| **Illness complications (multi-choice)** | Have you had any of the following complications due to your illness? Select all that apply. | None; Pneumonia; Severe pneumonia leading to hospitalization; Respiratory failure; Septic shock; Multiple organ dysfunction or failure; Blood clots; Stroke; Other |

**Supplementary Table 3.**
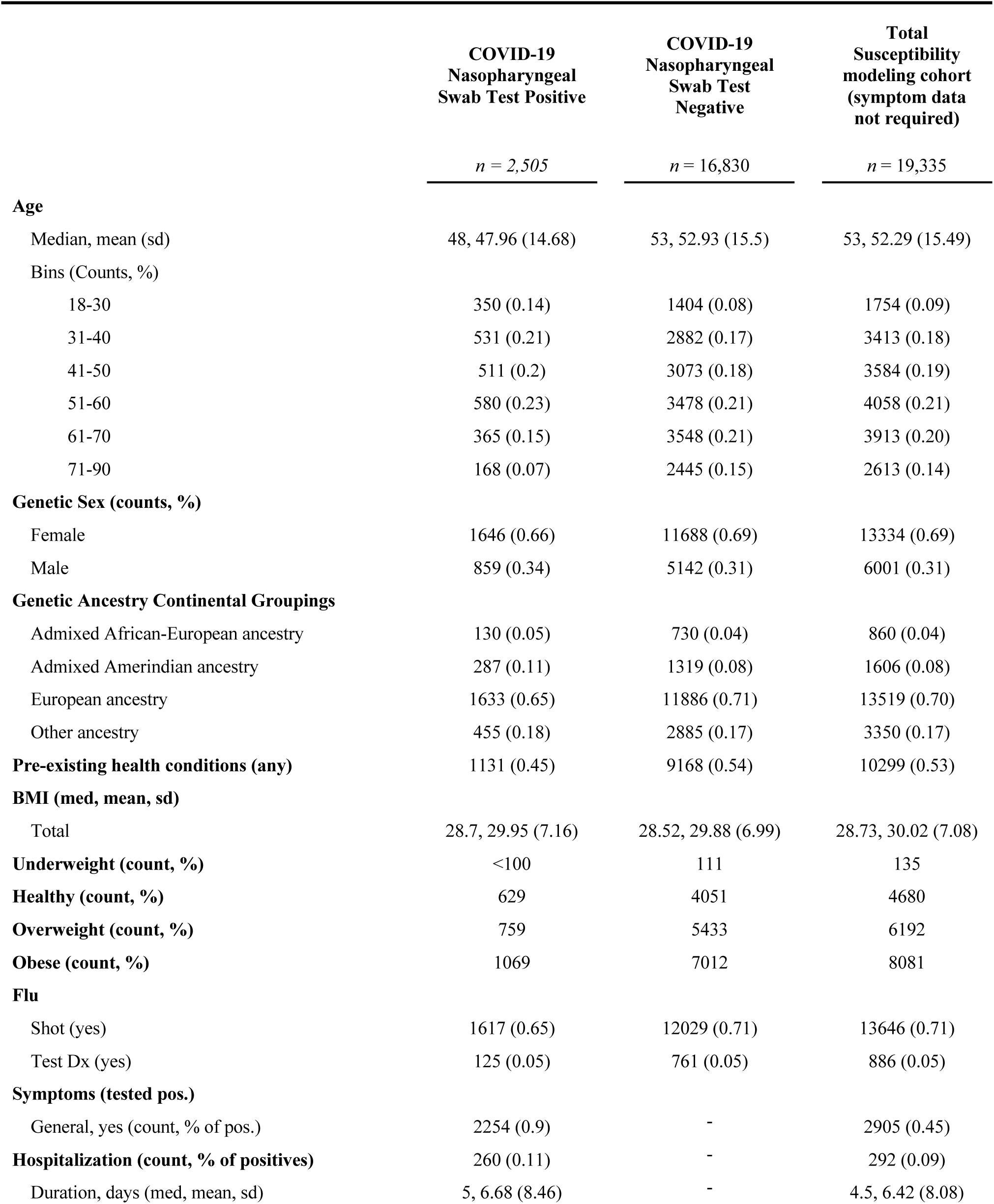
Demographics for susceptibility modeling cohort.

**Supplementary Table 4.** Demographics for severity modeling cohort.

|  | COVID-19<br>Nasopharyngeal<br>Swab Test Positive |
| --- | --- |
|  | *n* = 3,585 |
| **Age** |  |
| Median, mean (sd) | 49, 48.62 (14.68) |
| Bins (Counts, %) |  |
| 18-30 | 463 (0.13) |
| 31-40 | 725 (0.2) |
| 41-50 | 748 (0.21) |
| 51-60 | 832 (0.23) |
| 61-70 | 555 (0.15) |
| 71-90 | 262 (0.07) |
| **Genetic Sex (counts, %)** |  |
| Female | 2303 (0.64) |
| Male | 1282 (0.36) |
| **Genetic Ancestry Continental Groupings** |  |
| Admixed African-European ancestry | 203 (0.06) |
| Admixed Amerindian ancestry | 397 (0.11) |
| European ancestry | 2317 (0.65) |
| Other ancestry | 668 (0.19) |
| **Pre-existing health conditions (any)** | 1666 (0.46) |
| **BMI (med, mean, sd)** | 28.67, 29.83 (7.03) |
| **Underweight (count, %)** | <100 |
| **Healthy (count, %)** | 884 |
| **Overweight (count, %)** | 1128 |
| **Obese (count, %)** | 1498 |
| **Flu** |  |
| Shot (yes) | 2293 (0.64) |
| Test Dx (yes) | 174 (0.05) |
| **Symptoms (tested pos.)** |  |
| General, yes (count, % of pos.) | 3236 (0.9) |
| **Hospitalization (count, % of positives)** | 391 (0.11) |
| Duration, days (med, mean, sd) | 5, 7.34 (8.73) |

**Supplementary Table 5.** Demographics for susceptibility models with symptoms.

|  | Susceptibility<br>modeling cohort<br>(with symptom<br>data) |
| --- | --- |
|  | *n* = 4,094 |
| **Age** |  |
| Median, mean (sd) | 58, 55.7 (15.82) |
| Bins (Counts, %) |  |
| 18-30 | 293 (0.07) |
| 31-40 | 567 (0.14) |
| 41-50 | 641 (0.16) |
| 51-60 | 795 (0.19) |
| 61-70 | 982 (0.24) |
| 71-90 | 816 (0.20) |
| **Genetic Sex (counts, %)** |  |
| Female | 2744 (0.67) |
| Male | 1350 (0.33) |
| **Genetic Ancestry Continental Groupings** |  |
| Admixed African-European ancestry | 172 (0.04) |
| Admixed Amerindian ancestry | 298 (0.07) |
| European ancestry | 2927 (0.72) |
| Other ancestry | 666 (0.16) |
| **Pre-existing health conditions (any)** | 2205 (0.54) |
| **BMI (med, mean, sd)** | 28.59, 29.8 (6.84) |
| **Underweight (count, %)** | <100 |
| **Healthy (count, %)** | 1007 |
| **Overweight (count, %)** | 1335 |
| **Obese (count, %)** | 1662 |
| **Flu** |  |
| Shot (yes) | 2935 (0.72) |
| Test Dx (yes) | 160 (0.04) |
| **Symptoms (tested pos.)** |  |
| General, yes (count, % of pos.) | 1037 (0.25) |
| **Hospitalization (count, % of positives)** | <100 |
| Duration, days (med, mean, sd) | 5, 7.53 (10.11) |

**Supplementary Table 6.** Unadjusted risk factor odds ratios for COVID-19 susceptibility.

| RISK FACTOR | ODDS RATIO (OR) | OR 95% CI (BONFERRONI) | logOR | logOR S.E. | P-VALUE (NOMINAL) | P-VALUE (BONFERRONI) |
| --- | --- | --- | --- | --- | --- | --- |
| **AGE** |  |  |  |  |  |  |
| Ages 18-29 | 1.51 | (1.26, 1.81) | 0.4108 | 0.0563 | 3.05E-13 | 1.28E-11 |
| Ages 30-49 | 1.10 | (0.97, 1.24) | 0.0944 | 0.0377 | 1.22E-02 | 5.14E-01 |
| Ages 50-64 | 1.00 |  | 0 |  | nan | nan |
| Ages 65+ | 0.68 | (0.59, 0.79) | -0.3819 | 0.0454 | 0.00E+00 | 0.00E+00 |
| **GENETIC ANCESTRY** |  |  |  |  |  |  |
| Admixed African-European | 1.48 | (1.18, 1.85) | 0.3925 | 0.0694 | 1.57E-08 | 6.58E-07 |
| European | 1.00 |  | 0 |  | nan | nan |
| Admixed Amerindian | 1.49 | (1.26, 1.77) | 0.3995 | 0.0523 | 2.10E-14 | 8.82E-13 |
| Other | 1.21 | (1.06, 1.38) | 0.188 | 0.041 | 4.62E-06 | 1.94E-04 |
| **EXPOSURE** |  |  |  |  |  |  |
| Any | 8.72 | (7.42, 10.26) | 2.1659 | 0.0501 | 0.00E+00 | 0.00E+00 |
| Biological relative tested positive | 5.77 | (4.99, 6.68) | 1.7529 | 0.045 | 0.00E+00 | 0.00E+00 |
| Directly exposed to someone who tested positive | 6.94 | (6.02, 7.99) | 1.9366 | 0.0438 | 0.00E+00 | 0.00E+00 |
| Healthcare worker directly exposed | 1.44 | (1.26, 1.65) | 0.367 | 0.0408 | 2.31E-19 | 9.69E-18 |
| Household member tested positive | 26.03 | (22.26, 30.43) | 3.2591 | 0.0482 | 0.00E+00 | 0.00E+00 |
| **HEALTH CONDITIONS** |  |  |  |  |  |  |
| Asthma | 0.81 | (0.71, 0.94) | -0.2046 | 0.0446 | 4.55E-06 | 1.91E-04 |
| COPD (Chronic Obstructive Pulmonary Disease) | 0.58 | (0.41, 0.82) | -0.5374 | 0.1057 | 3.72E-07 | 1.56E-05 |
| Cancer treated in the past year | 0.53 | (0.36, 0.80) | -0.6283 | 0.1237 | 3.80E-07 | 1.59E-05 |
| Cardiovascular disease | 0.71 | (0.56, 0.91) | -0.3383 | 0.0762 | 9.12E-06 | 3.83E-04 |
| Chronic kidney disease | 0.54 | (0.33, 0.87) | -0.6241 | 0.148 | 2.48E-05 | 1.04E-03 |
| Diabetes | 0.86 | (0.71, 1.03) | -0.1547 | 0.0561 | 5.79E-03 | 2.43E-01 |
| Hypertension | 0.78 | (0.68, 0.89) | -0.2522 | 0.0411 | 8.08E-10 | 3.40E-08 |
| Other | 0.91 | (0.73, 1.13) | -0.0939 | 0.0675 | 1.64E-01 | 1.00E+00 |
| Other autoimmune disease | 0.77 | (0.65, 0.92) | -0.2571 | 0.054 | 1.95E-06 | 8.20E-05 |
| Other immunodeficiency disorder | 0.77 | (0.53, 1.14) | -0.2552 | 0.1194 | 3.25E-02 | 1.00E+00 |
| Other lung condition | 0.59 | (0.42, 0.82) | -0.5355 | 0.1037 | 2.45E-07 | 1.03E-05 |
| Any | 0.69 | (0.62, 0.77) | -0.3667 | 0.0336 | 0.00E+00 | 0.00E+00 |
| **OBESITY** |  |  |  |  |  |  |
| obesity I (BMI >= 30 and BMI < 35) | 0.99 | (0.86, 1.13) | -0.0146 | 0.043 | 7.35E-01 | 1.00E+00 |
| obesity II (BMI >= 35 and BMI < 40) | 1.00 | (0.83, 1.19) | -0.0042 | 0.056 | 9.41E-01 | 1.00E+00 |
| obesity III (BMI >= 40) | 0.97 | (0.80, 1.19) | -0.0255 | 0.0622 | 6.81E-01 | 1.00E+00 |
| Not obese | 1.00 |  | 0 |  | nan | nan |
| **GENETIC SEX** |  |  |  |  |  |  |
| Female | 1.00 |  | 0 |  | nan | nan |
| Male | 1.27 | (1.14, 1.42) | 0.2419 | 0.0329 | 1.89E-13 | 7.95E-12 |
| **SYMPTOMS** |  |  |  |  |  |  |
| Abdominal pain | 1.01 | (0.77, 1.33) | 0.0139 | 0.0841 | 8.69E-01 | 1.00E+00 |
| Body aches | 1.34 | (1.06, 1.69) | 0.2945 | 0.0717 | 4.04E-05 | 1.70E-03 |
| Change in taste or smell | 7.26 | (5.54, 9.50) | 1.9819 | 0.0832 | 1.62E-125 | 6.81E-124 |
| Chills | 1.37 | (1.09, 1.71) | 0.3126 | 0.0697 | 7.23E-06 | 3.04E-04 |
| Cough producing phlegm | 0.88 | (0.69, 1.12) | -0.1318 | 0.0746 | 7.74E-02 | 1.00E+00 |
| Diarrhea | 1.22 | (0.96, 1.56) | 0.2006 | 0.0745 | 7.11E-03 | 2.99E-01 |
| Dry cough | 1.12 | (0.89, 1.40) | 0.109 | 0.0694 | 1.16E-01 | 1.00E+00 |
| Feeling tired or fatigue | 1.41 | (1.05, 1.89) | 0.343 | 0.0901 | 1.41E-04 | 5.91E-03 |
| Fever | 1.60 | (1.28, 2.01) | 0.4715 | 0.0701 | 1.74E-11 | 7.30E-10 |
| Headache | 1.12 | (0.88, 1.41) | 0.1098 | 0.0728 | 1.32E-01 | 1.00E+00 |
| Nasal congestion | 0.74 | (0.59, 0.92) | -0.3072 | 0.0696 | 1.01E-05 | 4.24E-04 |
| Nausea or vomiting | 0.99 | (0.75, 1.32) | -0.0055 | 0.0867 | 9.50E-01 | 1.00E+00 |
| Runny nose | 0.59 | (0.47, 0.75) | -0.5245 | 0.0725 | 4.49E-13 | 1.89E-11 |
| Shortness of breath | 1.14 | (0.91, 1.44) | 0.1343 | 0.0705 | 5.67E-02 | 1.00E+00 |
| Sore throat | 0.49 | (0.39, 0.62) | -0.7052 | 0.0717 | 8.10E-23 | 0.00E+00 |

Unadjusted odds ratios for a positive COVID-19 test were calculated among survey participants reporting a test result (positive or negative) based on the self-reported response to the following question: “Have you been swab tested for COVID-19, commonly referred to as coronavirus?” The following groups served as reference populations for the odds ratio calculations: European genetic ancestry (Genetic Ancestry), Ages 50-64 (Age), Female (Genetic Sex), and Not Obese (BMI < 30, Obesity). Note that symptoms odds ratios were calculated in a binary fashion among symptomatic customers only (Methods).

**Supplementary Table 7.**
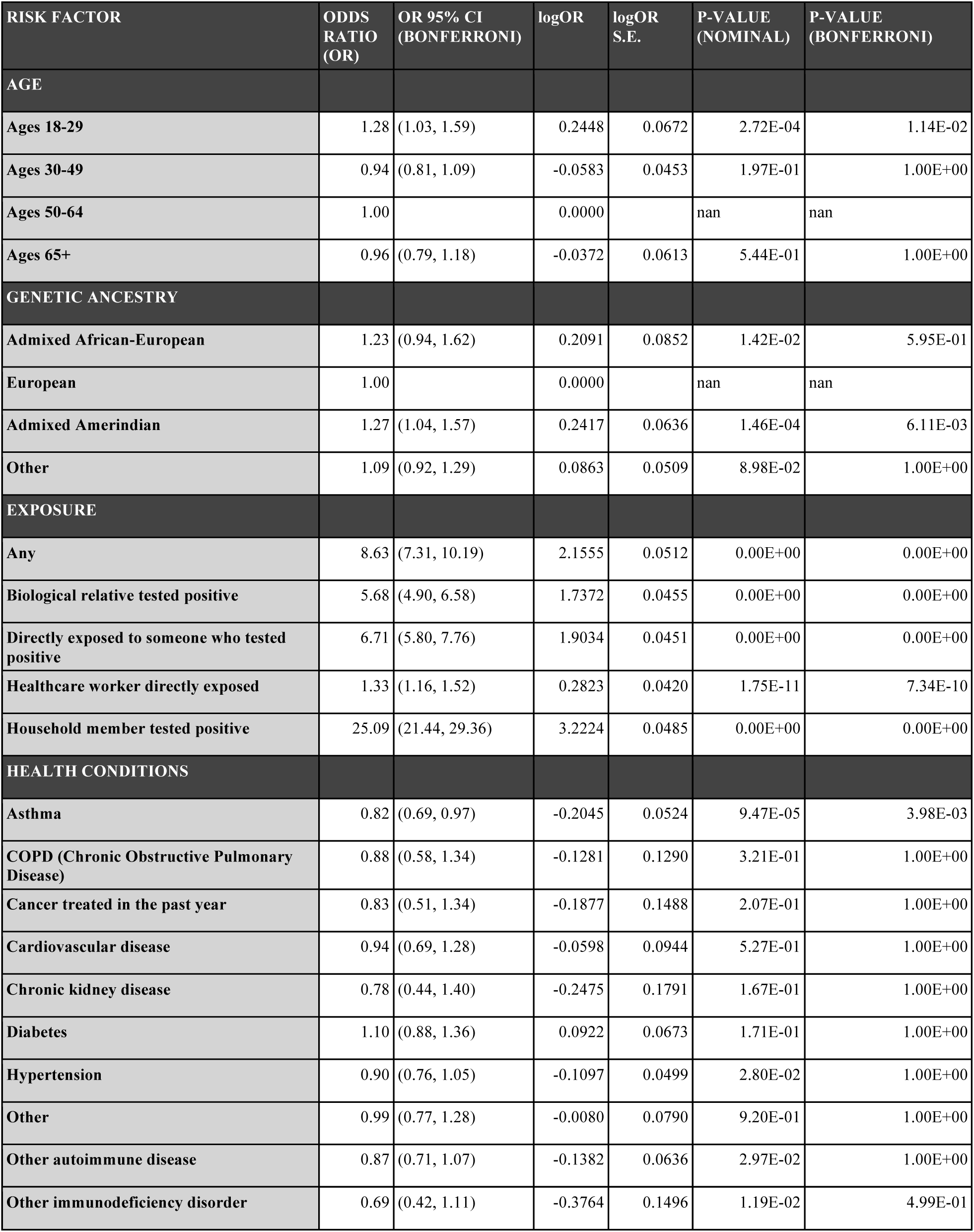
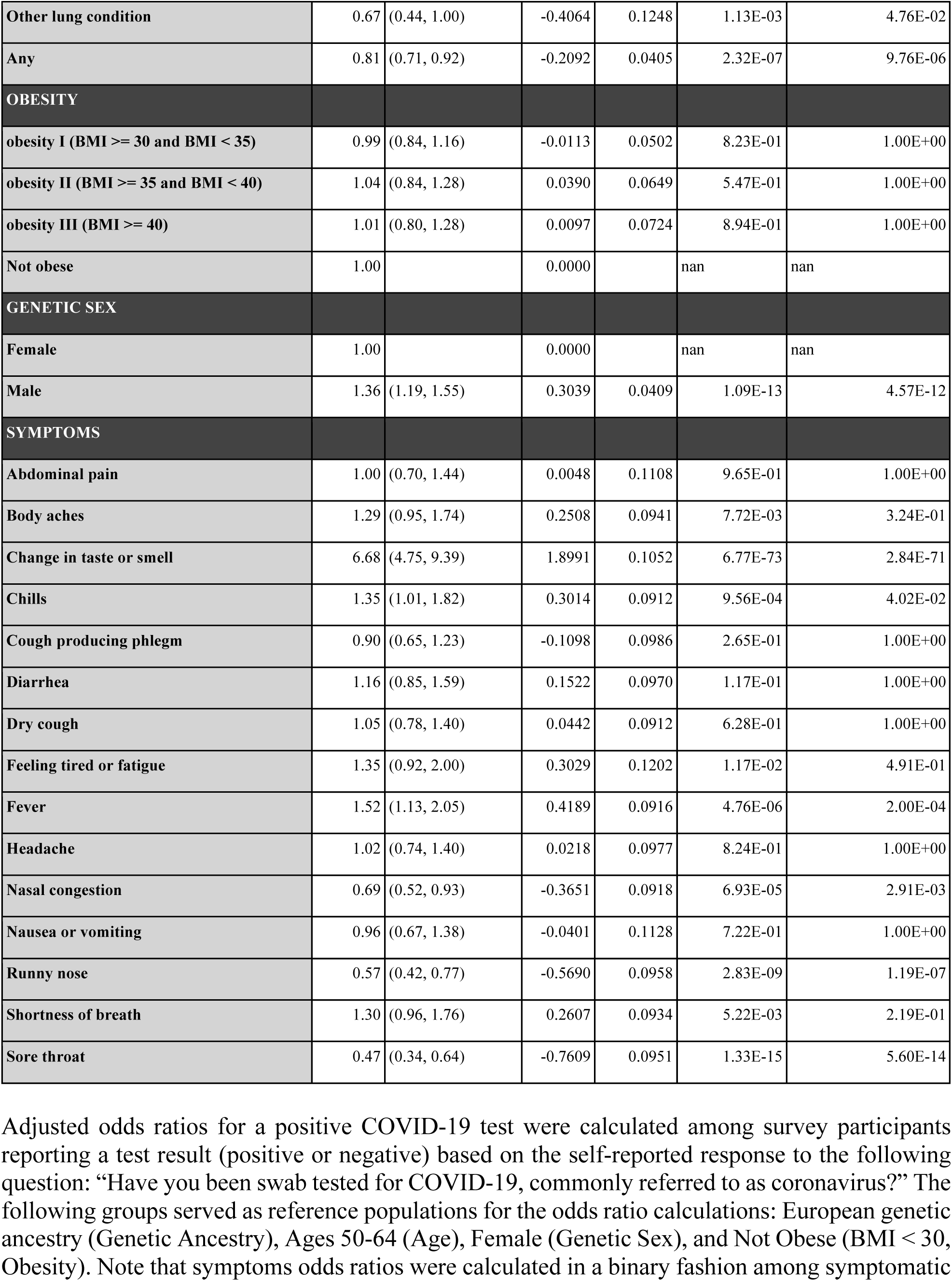
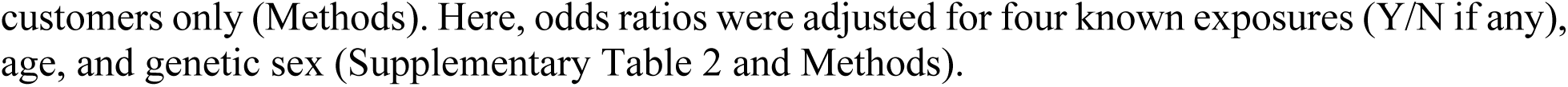
Risk factor odds ratios for COVID-19 susceptibility, adjusted for exposure, age, and sex.

**Supplementary Table 8.** Unadjusted risk factor odds ratios for a known “direct” COVID-19 exposure.

| RISK FACTOR | ODDS RATIO (OR) | OR 95% CI (BONFERRONI) | logOR | logOR S.E. | P-VALUE (NOMINAL) | P-VALUE (BONFERRONI) |
| --- | --- | --- | --- | --- | --- | --- |
| **AGE** |  |  |  |  |  |  |
| Ages 18-29 | 1.80 | (1.54, 2.12) | 0.5896 | 0.0519 | 6.64E-30 | 1.66E-28 |
| Ages 30-49 | 1.45 | (1.31, 1.60) | 0.3725 | 0.0325 | 2.00E-30 | 5.00E-29 |
| Ages 50-64 | 1.00 |  | 0.0000 |  | nan | nan |
| Ages 65+ | 0.32 | (0.28, 0.37) | -1.1427 | 0.0437 | 0.00E+00 | 0.00E+00 |
| **GENETIC ANCESTRY** |  |  |  |  |  |  |
| Admixed African-European | 1.49 | (1.23, 1.82) | 0.4012 | 0.0639 | 3.32E-10 | 8.31E-09 |
| European | 1.00 |  | 0.0000 |  | nan | nan |
| Admixed Amerindian | 1.40 | (1.21, 1.62) | 0.3384 | 0.0476 | 1.13E-12 | 2.82E-11 |
| Other | 1.17 | (1.05, 1.31) | 0.1584 | 0.0365 | 1.47E-05 | 3.67E-04 |
| **EXPOSURE** |  |  |  |  |  |  |
| Biological relative tested positive | 4.43 | (3.79, 5.18) | 1.4888 | 0.0503 | 1.39E-192 | 3.48E-191 |
| Healthcare worker directly exposed | 11.53 | (10.16, 13.08) | 2.4450 | 0.0408 | 0.00E+00 | 0.00E+00 |
| Household member tested positive | 22.53 | (17.88, 28.38) | 3.1148 | 0.0747 | 0.00E+00 | 0.00E+00 |
| **HEALTH CONDITIONS** |  |  |  |  |  |  |
| Asthma | 1.02 | (0.92, 1.14) | 0.0216 | 0.0353 | 5.40E-01 | 1.00E+00 |
| COPD (Chronic Obstructive Pulmonary Disease) | 0.44 | (0.33, 0.57) | -0.8297 | 0.0875 | 0.00E+00 | 0.00E+00 |
| Cancer treated in the past year | 0.36 | (0.26, 0.49) | -1.0310 | 0.1012 | 0.00E+00 | 0.00E+00 |
| Cardiovascular disease | 0.50 | (0.41, 0.61) | -0.6880 | 0.0647 | 0.00E+00 | 0.00E+00 |
| Chronic kidney disease | 0.39 | (0.27, 0.57) | -0.9295 | 0.1182 | 3.77E-15 | 9.44E-14 |
| Diabetes | 0.63 | (0.54, 0.73) | -0.4656 | 0.0473 | 0.00E+00 | 0.00E+00 |
| Hypertension | 0.73 | (0.66, 0.81) | -0.3181 | 0.0332 | 0.00E+00 | 0.00E+00 |
| Other | 0.99 | (0.84, 1.18) | -0.0080 | 0.0556 | 8.86E-01 | 1.00E+00 |
| Other autoimmune disease | 0.87 | (0.76, 0.99) | -0.1421 | 0.0424 | 8.06E-04 | 2.02E-02 |
| Other immunodeficiency disorder | 0.87 | (0.65, 1.17) | -0.1389 | 0.0953 | 1.45E-01 | 1.00E+00 |
| Other lung condition | 0.62 | (0.48, 0.78) | -0.4859 | 0.0778 | 4.22E-10 | 1.05E-08 |
| Any | 0.71 | (0.65, 0.78) | -0.3393 | 0.0276 | 0.00E+00 | 0.00E+00 |

| OBESITY |  |  |  |  |  |  |
| --- | --- | --- | --- | --- | --- | --- |
| obesity I (BMI ≥ 30 and BMI < 35) | 1.03 | (0.93, 1.15) | 0.0338 | 0.0350 | 3.35E-01 | 1.00E+00 |
| obesity II (BMI ≥ 35 and BMI < 40) | 1.03 | (0.90, 1.19) | 0.0343 | 0.0456 | 4.52E-01 | 1.00E+00 |
| obesity III (BMI ≥ 40) | 1.10 | (0.94, 1.28) | 0.0927 | 0.0499 | 6.31E-02 | 1.00E+00 |
| Not obese | 1.00 |  | 0.0000 |  | nan | nan |
| GENETIC SEX |  |  |  |  |  |  |
| Female | 1.00 |  | 0.0000 |  | nan | nan |
| Male | 0.96 | (0.87, 1.05) | -0.0443 | 0.0295 | 1.33E-01 | 1.00E+00 |

Unadjusted odds ratios for a known “direct” exposure to COVID-19 were calculated based on the self-reported response to the following question: “Have you been directly exposed to someone who has tested positive for COVID-19?” The following groups served as reference populations for the odds ratio calculations: European genetic ancestry (Genetic Ancestry), Ages 50-64 (Age), Female (Genetic Sex), and Not Obese (BMI < 30, Obesity).

**Supplementary Table 9.**
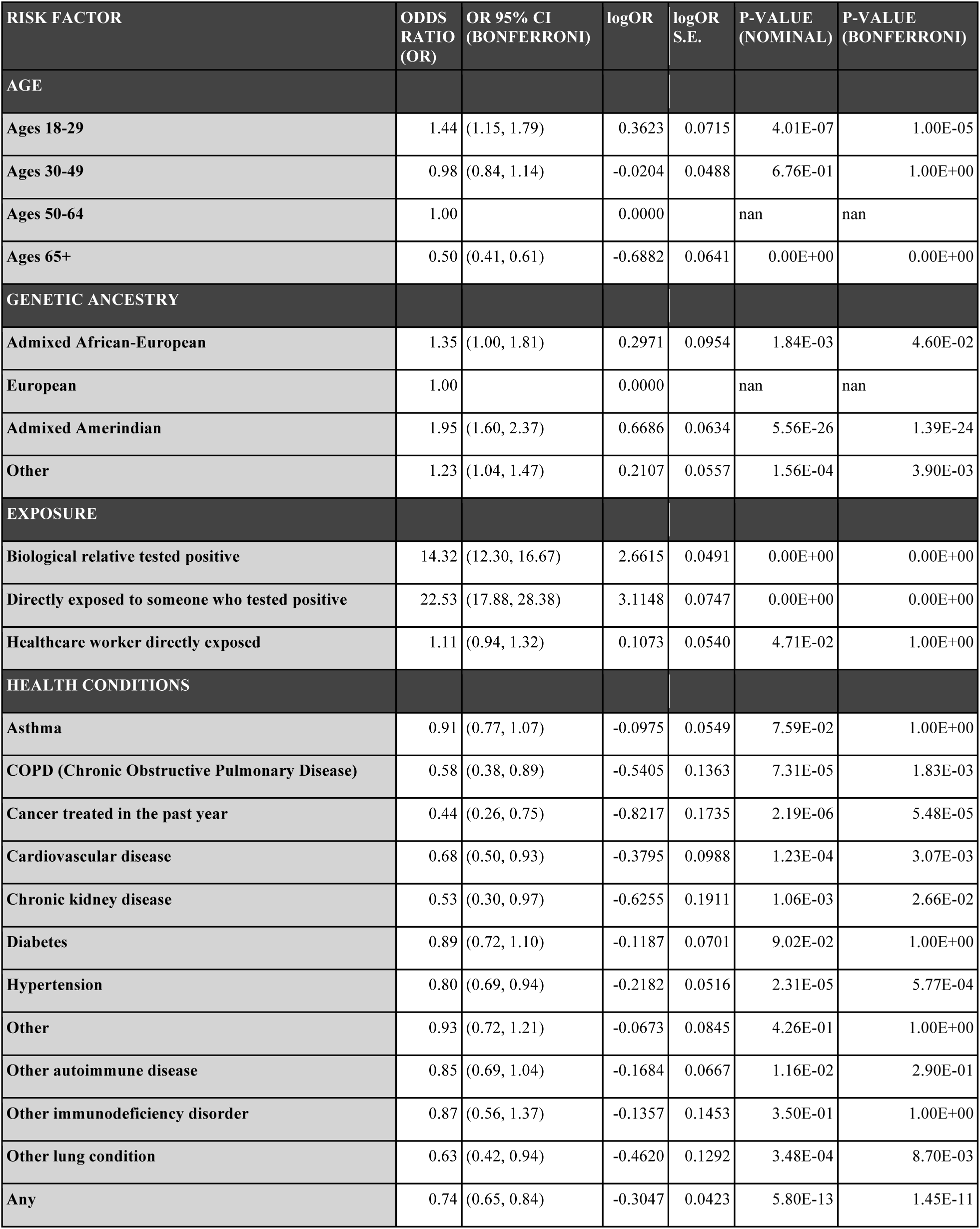
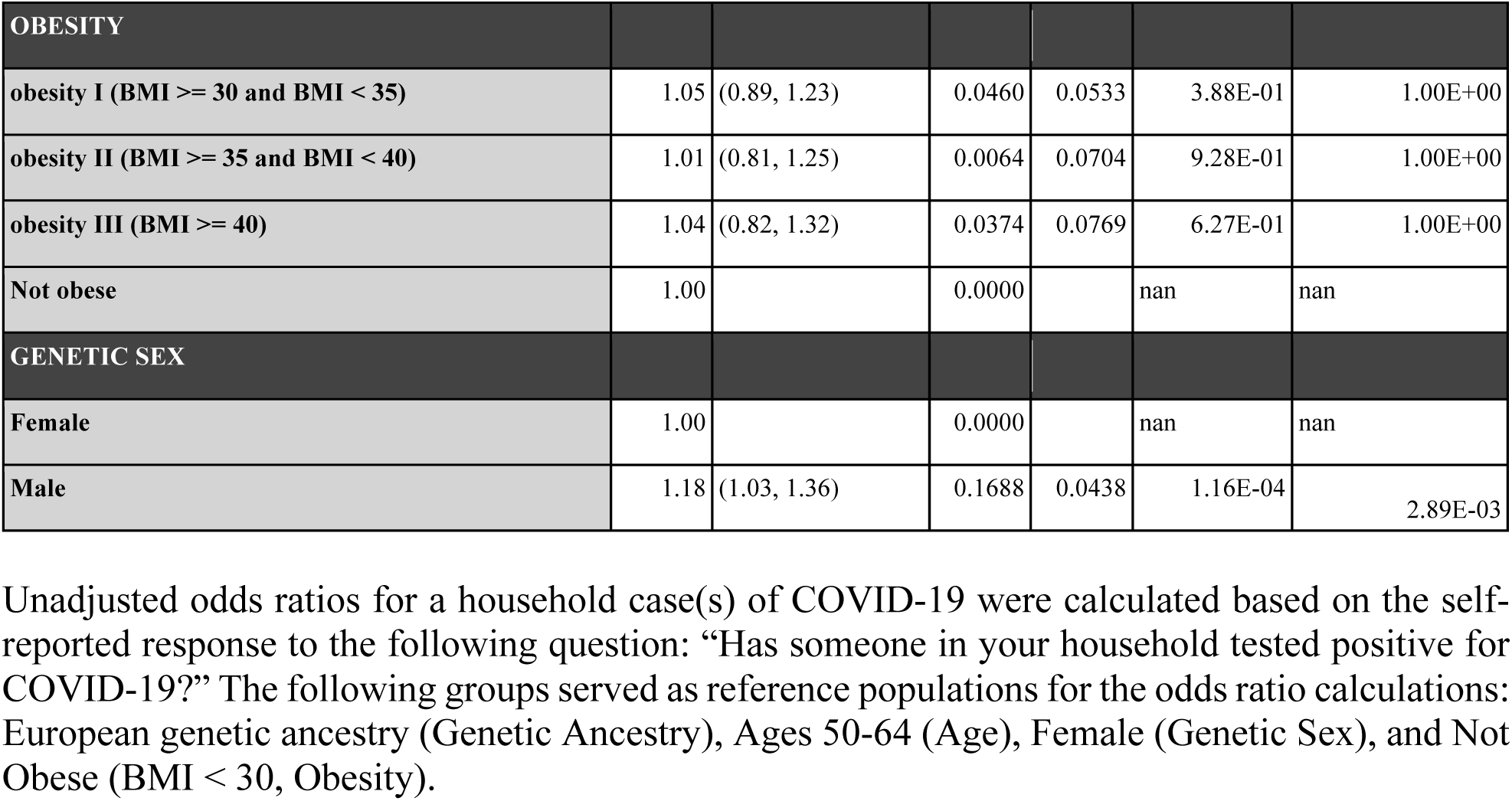
Unadjusted risk factor odds ratios for a household case(s) of COVID-19.

**Supplementary Table 10.** Unadjusted risk factor odds ratios for a COVID-19 case(s) amongst close biological relatives (parent, grandparent, child, or sibling)

| RISK FACTOR | ODDS RATIO (OR) | OR 95% CI (BONFERRONI) | logOR | logOR S.E. | P-VALUE (NOMINAL) | P-VALUE (BONFERRONI) |
| --- | --- | --- | --- | --- | --- | --- |
| **AGE** |  |  |  |  |  |  |
| Ages 18-29 | 1.48 | (1.18, 1.86) | 0.3934 | 0.0730 | 7.16E-08 | 1.79E-06 |
| Ages 30-49 | 1.02 | (0.88, 1.19) | 0.0222 | 0.0502 | 6.59E-01 | 1.00E+00 |
| Ages 50-64 | 1.00 |  | 0.0000 |  | nan | nan |
| Ages 65+ | 0.70 | (0.58, 0.85) | -0.3523 | 0.0605 | 5.90E-09 | 1.47E-07 |
| **GENETIC ANCESTRY** |  |  |  |  |  |  |
| Admixed African-European | 1.69 | (1.28, 2.22) | 0.5221 | 0.0896 | 5.62E-09 | 1.41E-07 |
| European | 1.00 |  | 0.0000 |  | nan | nan |
| Admixed Amerindian | 2.08 | (1.71, 2.53) | 0.7333 | 0.0634 | 5.85E-31 | 1.46E-29 |
| Other | 1.42 | (1.20, 1.68) | 0.3472 | 0.0547 | 2.14E-10 | 5.34E-09 |
| **EXPOSURE** |  |  |  |  |  |  |
| Directly exposed to someone who tested positive | 4.43 | (3.79, 5.18) | 1.4888 | 0.0503 | 1.39E-192 | 3.48E-191 |
| Healthcare worker directly exposed | 1.04 | (0.87, 1.23) | 0.0384 | 0.0558 | 4.91E-01 | 1.00E+00 |
| Household member tested positive | 14.32 | (12.30, 16.67) | 2.6615 | 0.0491 | 0.00E+00 | 0.00E+00 |
| **HEALTH CONDITIONS** |  |  |  |  |  |  |
| Asthma | 0.98 | (0.82, 1.16) | -0.0245 | 0.0547 | 6.54E-01 | 1.00E+00 |
| COPD (Chronic Obstructive Pulmonary Disease) | 0.68 | (0.46, 1.02) | -0.3791 | 0.1293 | 3.37E-03 | 8.42E-02 |
| Cancer treated in the past year | 0.60 | (0.37, 0.97) | -0.5105 | 0.1542 | 9.33E-04 | 2.33E-02 |
| Cardiovascular disease | 0.77 | (0.57, 1.04) | -0.2600 | 0.0959 | 6.72E-03 | 1.68E-01 |
| Chronic kidney disease | 0.52 | (0.28, 0.95) | -0.6619 | 0.1977 | 8.12E-04 | 2.03E-02 |
| Diabetes | 1.04 | (0.84, 1.28) | 0.0368 | 0.0677 | 5.86E-01 | 1.00E+00 |
| Hypertension | 0.88 | (0.75, 1.03) | -0.1281 | 0.0513 | 1.25E-02 | 3.12E-01 |
| Other | 1.03 | (0.80, 1.33) | 0.0297 | 0.0829 | 7.20E-01 | 1.00E+00 |
| Other autoimmune disease | 0.90 | (0.73, 1.10) | -0.1057 | 0.0663 | 1.11E-01 | 1.00E+00 |
| Other immunodeficiency disorder | 0.99 | (0.65, 1.53) | -0.0065 | 0.1394 | 9.63E-01 | 1.00E+00 |
| Other lung condition | 0.96 | (0.69, 1.36) | -0.0368 | 0.1104 | 7.39E-01 | 1.00E+00 |
| Any | 0.85 | (0.74, 0.97) | -0.1668 | 0.0429 | 1.02E-04 | 2.55E-03 |

| OBESITY |  |  |  |  |  |  |
| --- | --- | --- | --- | --- | --- | --- |
| obesity I (BMI ≥ 30 and BMI < 35) | 1.03 | (0.87, 1.22) | 0.0327 | 0.0541 | 5.45E-01 | 1.00E+00 |
| obesity II (BMI ≥ 35 and BMI < 40) | 0.88 | (0.70, 1.11) | -0.1292 | 0.0748 | 8.41E-02 | 1.00E+00 |
| obesity III (BMI ≥ 40) | 1.10 | (0.87, 1.39) | 0.0945 | 0.0761 | 2.14E-01 | 1.00E+00 |
| Not obese | 1.00 |  | 0.0000 |  | nan | nan |
| GENETIC SEX |  |  |  |  |  |  |
| Female | 1.00 |  | 0.0000 |  | nan | nan |
| Male | 0.87 | (0.75, 1.00) | -0.1404 | 0.0462 | 2.39E-03 | 5.99E-02 |

Unadjusted odds ratios for a case(s) of COVID-19 among close biological relatives (grandparent, parent, child, or sibling) were calculated based on the self-reported response to the following question: “Have any of your biological relatives tested positive for COVID-19? Select all that apply.” The following groups served as reference populations for the odds ratio calculations: European genetic ancestry (Genetic Ancestry), Ages 50-64 (Age), Female (Genetic Sex), and Not Obese (BMI < 30, Obesity).

**Supplementary Table 11.** Unadjusted risk factor odds ratios for COVID-19 hospitalization.

| RISK FACTOR | ODDS RATIO (OR) | OR 95% CI (BONFERRONI) | logOR | logOR S.E. | P-VALUE (NOMINAL) | P-VALUE (BONFERRONI) |
| --- | --- | --- | --- | --- | --- | --- |
| **AGE** |  |  |  |  |  |  |
| Ages 18-29 | 0.42 | (0.21, 0.83) | -0.8793 | 0.2143 | 4.06E-05 | 1.71E-03 |
| Ages 30-49 | 0.54 | (0.36, 0.81) | -0.6143 | 0.1246 | 8.24E-07 | 3.46E-05 |
| Ages 50-64 | 1.00 |  | 0.0000 |  | nan | nan |
| Ages 65+ | 1.70 | (1.13, 2.56) | 0.5325 | 0.1263 | 2.49E-05 | 1.05E-03 |
| **GENETIC ANCESTRY** |  |  |  |  |  |  |
| Admixed African-European | 1.44 | (0.78, 2.65) | 0.3625 | 0.1884 | 5.43E-02 | 1.00E+00 |
| European | 1.00 |  | 0.0000 |  | nan | nan |
| Admixed Amerindian | 0.77 | (0.44, 1.36) | -0.2579 | 0.1745 | 1.39E-01 | 1.00E+00 |
| Other | 0.91 | (0.59, 1.40) | -0.0964 | 0.1335 | 4.70E-01 | 1.00E+00 |
| **EXPOSURE** |  |  |  |  |  |  |
| Any | 0.74 | (0.46, 1.19) | -0.3048 | 0.1483 | 3.98E-02 | 1.00E+00 |
| Biological relative tested positive | 1.17 | (0.81, 1.69) | 0.1550 | 0.1136 | 1.73E-01 | 1.00E+00 |
| Directly exposed to someone who tested positive | 0.63 | (0.42, 0.96) | -0.4595 | 0.1279 | 3.28E-04 | 1.38E-02 |
| Healthcare worker directly exposed | 0.53 | (0.34, 0.84) | -0.6317 | 0.1423 | 9.09E-06 | 3.82E-04 |
| Household member tested positive | 0.91 | (0.65, 1.27) | -0.0971 | 0.1033 | 3.47E-01 | 1.00E+00 |
| **HEALTH CONDITIONS** |  |  |  |  |  |  |
| Asthma | 1.42 | (0.95, 2.13) | 0.3525 | 0.1249 | 4.77E-03 | 2.00E-01 |
| COPD (Chronic Obstructive Pulmonary Disease) | 3.48 | (1.65, 7.32) | 1.2469 | 0.2296 | 5.64E-08 | 2.37E-06 |
| Cancer treated in the past year | 3.13 | (1.29, 7.57) | 1.1396 | 0.2730 | 2.99E-05 | 1.26E-03 |
| Cardiovascular disease | 2.82 | (1.61, 4.96) | 1.0374 | 0.1740 | 2.50E-09 | 1.05E-07 |
| Chronic kidney disease | 8.74 | (3.44, 22.21) | 2.1684 | 0.2876 | 4.71E-14 | 1.98E-12 |
| Diabetes | 3.03 | (1.98, 4.63) | 1.1085 | 0.1311 | 2.78E-17 | 1.17E-15 |
| Hypertension | 2.01 | (1.41, 2.88) | 0.7002 | 0.1105 | 2.37E-10 | 9.96E-09 |
| Other | 1.65 | (0.93, 2.93) | 0.5038 | 0.1767 | 4.35E-03 | 1.83E-01 |
| Other autoimmune disease | 1.32 | (0.81, 2.17) | 0.2811 | 0.1523 | 6.49E-02 | 1.00E+00 |
| Other immunodeficiency disorder | 1.98 | (0.77, 5.09) | 0.6826 | 0.2915 | 1.92E-02 | 8.05E-01 |
| Other lung condition | 2.84 | (1.32, 6.10) | 1.0423 | 0.2364 | 1.04E-05 | 4.35E-04 |
| Any | 2.70 | (1.90, 3.83) | 0.9919 | 0.1081 | 4.43E-20 | 1.86E-18 |
| **OBESITY** |  |  |  |  |  |  |
| obesity I (BMI >= 30 and BMI < 35) | 1.41 | (0.94, 2.12) | 0.3418 | 0.1258 | 6.61E-03 | 2.78E-01 |
| obesity II (BMI >= 35 and BMI < 40) | 1.21 | (0.70, 2.09) | 0.1883 | 0.1693 | 2.66E-01 | 1.00E+00 |
| obesity III (BMI >= 40) | 1.62 | (0.93, 2.83) | 0.4820 | 0.1719 | 5.04E-03 | 2.12E-01 |
| Not obese | 1.00 |  | 0.0000 |  | nan | nan |
| **GENETIC SEX** |  |  |  |  |  |  |
| Female | 1.00 |  | 0.0000 |  | nan | nan |
| Male | 1.27 | (0.92, 1.77) | 0.2418 | 0.1015 | 1.72E-02 | 7.22E-01 |
| **SYMPTOMS** |  |  |  |  |  |  |
| Abdominal pain | 1.76 | (1.23, 2.52) | 0.5638 | 0.1112 | 3.92E-07 | 1.65E-05 |
| Body aches | 1.39 | (0.96, 2.02) | 0.3309 | 0.1142 | 3.75E-03 | 1.57E-01 |
| Change in taste or smell | 0.77 | (0.55, 1.07) | -0.2657 | 0.1039 | 1.05E-02 | 4.42E-01 |
| Chills | 1.82 | (1.29, 2.57) | 0.5991 | 0.1061 | 1.65E-08 | 6.92E-07 |
| Cough producing phlegm | 1.56 | (1.11, 2.20) | 0.4474 | 0.1054 | 2.19E-05 | 9.18E-04 |
| Diarrhea | 2.12 | (1.52, 2.95) | 0.7521 | 0.1021 | 1.75E-13 | 7.35E-12 |
| Dry cough | 2.59 | (1.81, 3.71) | 0.9528 | 0.1104 | 6.26E-18 | 2.63E-16 |
| Feeling tired or fatigue | 2.54 | (1.35, 4.78) | 0.9335 | 0.1949 | 1.68E-06 | 7.04E-05 |
| Fever | 3.33 | (2.26, 4.90) | 1.2032 | 0.1192 | 5.93E-24 | 2.49E-22 |
| Headache | 0.89 | (0.63, 1.26) | -0.1125 | 0.1063 | 2.90E-01 | 1.00E+00 |
| Nasal congestion | 0.94 | (0.67, 1.31) | -0.0610 | 0.1030 | 5.54E-01 | 1.00E+00 |
| Nausea or vomiting | 2.26 | (1.58, 3.23) | 0.8156 | 0.1104 | 1.52E-13 | 6.39E-12 |
| Runny nose | 1.09 | (0.76, 1.56) | 0.0831 | 0.1118 | 4.57E-01 | 1.00E+00 |
| Shortness of breath | 7.52 | (4.92, 11.49) | 2.0172 | 0.1309 | 1.48E-53 | 6.21E-52 |
| Sore throat | 1.04 | (0.72, 1.50) | 0.0386 | 0.1127 | 7.32E-01 | 1.00E+00 |

Unadjusted odds ratios for hospitalization due to COVID-19 infection were calculated among
those reporting a positive COVID-19 test result. The following groups served as reference
populations for the odds ratio calculations: European genetic ancestry (Genetic Ancestry), Ages
50-64 (Age), Female (Genetic Sex), and Not Obese (BMI < 30, Obesity). Note that symptoms
odds ratios were calculated in a binary fashion among symptomatic customers only (Methods).

**Supplementary Table 12.** Risk factor odds ratios for COVID-19 hospitalization, adjusted for health conditions, obesity, age, and sex.

| RISK FACTOR | ODDS RATIO (OR) | OR 95% CI (BONFERRONI) | logOR | logOR S.E. | P-VALUE (NOMINAL) | P-VALUE (BONFERRONI) |
| --- | --- | --- | --- | --- | --- | --- |
| **AGE** |  |  |  |  |  |  |
| Ages 18-29 | 0.56 | (0.26, 1.20) | -0.5815 | 0.2359 | 1.37E-02 | 5.75E-01 |
| Ages 30-49 | 0.60 | (0.39, 0.92) | -0.5110 | 0.1333 | 1.26E-04 | 5.28E-03 |
| Ages 50-64 | 1.00 |  | 0.0000 |  | nan | nan |
| Ages 65+ | 1.60 | (1.02, 2.51) | 0.4713 | 0.1381 | 6.43E-04 | 2.70E-02 |
| **GENETIC ANCESTRY** |  |  |  |  |  |  |
| Admixed African-European | 1.57 | (0.81, 3.03) | 0.4513 | 0.2026 | 2.59E-02 | 1.00E+00 |
| European | 1.00 |  | 0.0000 |  | nan | nan |
| Admixed Amerindian | 1.11 | (0.59, 2.07) | 0.1035 | 0.1922 | 5.90E-01 | 1.00E+00 |
| Other | 0.95 | (0.59, 1.54) | -0.0493 | 0.1490 | 7.41E-01 | 1.00E+00 |
| **EXPOSURE** |  |  |  |  |  |  |
| Any | 0.81 | (0.48, 1.39) | -0.2060 | 0.1653 | 2.13E-01 | 1.00E+00 |
| Biological relative tested positive | 1.16 | (0.78, 1.74) | 0.1498 | 0.1248 | 2.30E-01 | 1.00E+00 |
| Directly exposed to someone who tested positive | 0.68 | (0.43, 1.07) | -0.3870 | 0.1397 | 5.61E-03 | 2.36E-01 |
| Healthcare worker directly exposed | 0.63 | (0.38, 1.04) | -0.4685 | 0.1567 | 2.79E-03 | 1.17E-01 |
| Household member tested positive | 0.88 | (0.62, 1.26) | -0.1245 | 0.1109 | 2.62E-01 | 1.00E+00 |
| **HEALTH CONDITIONS** |  |  |  |  |  |  |
| Asthma | 1.42 | (0.92, 2.20) | 0.3537 | 0.1343 | 8.42E-03 | 3.54E-01 |
| COPD (Chronic Obstructive Pulmonary Disease) | 2.25 | (0.99, 5.14) | 0.8123 | 0.2542 | 1.40E-03 | 5.87E-02 |
| Cancer treated in the past year | 1.90 | (0.72, 5.04) | 0.6422 | 0.3012 | 3.30E-02 | 1.00E+00 |
| Cardiovascular disease | 1.67 | (0.90, 3.11) | 0.5139 | 0.1914 | 7.24E-03 | 3.04E-01 |
| Chronic kidney disease | 5.62 | (2.10, 15.03) | 1.7267 | 0.3034 | 1.26E-08 | 5.30E-07 |
| Diabetes | 2.22 | (1.39, 3.55) | 0.7988 | 0.1442 | 3.06E-08 | 1.28E-06 |
| Hypertension | 1.32 | (0.88, 1.97) | 0.2757 | 0.1239 | 2.61E-02 | 1.00E+00 |
| Other | 1.47 | (0.80, 2.71) | 0.3878 | 0.1875 | 3.86E-02 | 1.00E+00 |
| Other autoimmune disease | 1.41 | (0.83, 2.40) | 0.3461 | 0.1635 | 3.42E-02 | 1.00E+00 |
| Other immunodeficiency disorder | 1.83 | (0.68, 4.93) | 0.6032 | 0.3062 | 4.88E-02 | 1.00E+00 |
| **Other lung condition** | 1.87 | (0.83, 4.22) | 0.6242 | 0.2514 | 1.30E-02 | 5.47E-01 |
| **Any** | 2.08 | (1.42, 3.04) | 0.7333 | 0.1172 | 3.93E-10 | 1.65E-08 |
| **OBEESITY** |  |  |  |  |  |  |
| **obesity I (BMI >= 30 and BMI < 35)** | 1.19 | (0.77, 1.82) | 0.1707 | 0.1323 | 1.97E-01 | 1.00E+00 |
| **obesity II (BMI >= 35 and BMI < 40)** | 1.13 | (0.64, 2.00) | 0.1249 | 0.1750 | 4.75E-01 | 1.00E+00 |
| **obesity III (BMI >= 40)** | 1.59 | (0.88, 2.87) | 0.4631 | 0.1821 | 1.10E-02 | 4.62E-01 |
| **Not obese** | 1.00 |  | 0.0000 |  | nan | nan |
| **GENETIC SEX** |  |  |  |  |  |  |
| **Female** | 1.00 |  | 0.0000 |  | nan | nan |
| **Male** | 1.22 | (0.85, 1.74) | 0.1960 | 0.1103 | 7.57E-02 | 1.00E+00 |
| **SYMPTOMS** |  |  |  |  |  |  |
| **Abdominal pain** | 1.66 | (1.12, 2.46) | 0.5086 | 0.1214 | 2.81E-05 | 1.18E-03 |
| **Body aches** | 1.44 | (0.96, 2.15) | 0.3646 | 0.1240 | 3.27E-03 | 1.37E-01 |
| **Change in taste or smell** | 0.89 | (0.62, 1.29) | -0.1159 | 0.1139 | 3.09E-01 | 1.00E+00 |
| **Chills** | 1.94 | (1.33, 2.82) | 0.6609 | 0.1156 | 1.07E-08 | 4.51E-07 |
| **Cough producing phlegm** | 1.41 | (0.97, 2.05) | 0.3470 | 0.1150 | 2.54E-03 | 1.07E-01 |
| **Diarrhea** | 2.23 | (1.55, 3.20) | 0.8014 | 0.1120 | 8.21E-13 | 3.45E-11 |
| **Dry cough** | 2.49 | (1.69, 3.67) | 0.9114 | 0.1196 | 2.55E-14 | 1.07E-12 |
| **Feeling tired or fatigue** | 2.81 | (1.38, 5.74) | 1.0333 | 0.2203 | 2.73E-06 | 1.15E-04 |
| **Fever** | 3.49 | (2.29, 5.34) | 1.2510 | 0.1309 | 1.24E-21 | 5.19E-20 |
| **Headache** | 1.05 | (0.72, 1.54) | 0.0487 | 0.1180 | 6.80E-01 | 1.00E+00 |
| **Nasal congestion** | 0.93 | (0.65, 1.34) | -0.0698 | 0.1122 | 5.34E-01 | 1.00E+00 |
| **Nausea or vomiting** | 2.41 | (1.62, 3.59) | 0.8801 | 0.1230 | 8.38E-13 | 3.52E-11 |
| **Runny nose** | 0.93 | (0.62, 1.39) | -0.0710 | 0.1232 | 5.65E-01 | 1.00E+00 |
| **Shortness of breath** | 8.20 | (5.14, 13.08) | 2.1042 | 0.1440 | 2.37E-48 | 9.96E-47 |
| **Sore throat** | 1.11 | (0.75, 1.66) | 0.1074 | 0.1224 | 3.80E-01 | 1.00E+00 |

Adjusted odds ratios for hospitalization due to COVID-19 infection were calculated among those reporting a positive COVID-19 test result. The following groups served as reference populations for the odds ratio calculations: European genetic ancestry (Genetic Ancestry), Ages 50-64 (Age), Female (Genetic Sex), and Not Obese (BMI < 30, Obesity). Note that symptoms odds ratios were calculated in a binary fashion among symptomatic customers only (Methods). Here, odds ratios were adjusted for age, sex, comorbidities (Y/N if any), and obesity (Y/N if BMI > 30, Methods).

**Supplementary Table 13.** Unadjusted risk factor odds ratios for progression of COVID-19 to a critical case.

| RISK FACTOR | ODDS RATIO (OR) | OR 95% CI (BONFERRONI) | logOR | logOR S.E. | P-VALUE (NOMINAL) | P-VALUE (BONFERRONI) |
| --- | --- | --- | --- | --- | --- | --- |
| **AGE** |  |  |  |  |  |  |
| Ages 18-29 | 0.35 | (0.13, 0.95) | -1.0599 | 0.3114 | 6.64E-04 | 2.79E-02 |
| Ages 30-49 | 0.55 | (0.32, 0.94) | -0.5983 | 0.1652 | 2.92E-04 | 1.23E-02 |
| Ages 50-64 | 1 |  | 0 |  | nan | nan |
| Ages 65+ | 1.65 | (0.97, 2.80) | 0.5001 | 0.1632 | 2.19E-03 | 9.18E-02 |
| **GENETIC ANCESTRY** |  |  |  |  |  |  |
| Admixed African-European | 2.07 | (1.03, 4.17) | 0.7269 | 0.2166 | 7.91E-04 | 3.32E-02 |
| European | 1 |  | 0 |  | nan | nan |
| Admixed Amerindian | 0.75 | (0.35, 1.62) | -0.2871 | 0.2377 | 2.27E-01 | 1.00E+00 |
| Other | 0.82 | (0.45, 1.49) | -0.2044 | 0.1853 | 2.70E-01 | 1.00E+00 |
| **EXPOSURE** |  |  |  |  |  |  |
| Any | 0.77 | (0.40, 1.45) | -0.2675 | 0.1981 | 1.77E-01 | 1.00E+00 |
| Biological relative tested positive | 1.15 | (0.70, 1.86) | 0.1356 | 0.1502 | 3.67E-01 | 1.00E+00 |
| Directly exposed to someone who tested positive | 0.59 | (0.34, 1.01) | -0.5348 | 0.1691 | 1.56E-03 | 6.55E-02 |
| Healthcare worker directly exposed | 0.53 | (0.29, 0.99) | -0.6277 | 0.1912 | 1.03E-03 | 4.31E-02 |
| Household member tested positive | 1.00 | (0.64, 1.55) | -0.0001 | 0.1353 | 1.00E+00 | 1.00E+00 |
| **HEALTH CONDITIONS** |  |  |  |  |  |  |
| Asthma | 1.26 | (0.74, 2.17) | 0.235 | 0.1672 | 1.60E-01 | 1.00E+00 |
| COPD (Chronic Obstructive Pulmonary Disease) | 3.76 | (1.57, 9.02) | 1.3257 | 0.2696 | 8.79E-07 | 3.69E-05 |
| Cancer treated in the past year | 2.07 | (0.60, 7.12) | 0.7266 | 0.3814 | 5.68E-02 | 1.00E+00 |
| Cardiovascular disease | 2.44 | (1.17, 5.05) | 0.8906 | 0.2251 | 7.60E-05 | 3.19E-03 |
| Chronic kidney disease | 8.00 | (2.92, 21.91) | 2.0794 | 0.3109 | 2.26E-11 | 9.47E-10 |
| Diabetes | 3.14 | (1.85, 5.32) | 1.1431 | 0.1629 | 2.29E-12 | 9.61E-11 |
| Hypertension | 2.06 | (1.30, 3.27) | 0.7214 | 0.1425 | 4.15E-07 | 1.74E-05 |
| Other | 1.07 | (0.45, 2.54) | 0.0649 | 0.2674 | 8.08E-01 | 1.00E+00 |
| Other autoimmune disease | 1.35 | (0.71, 2.55) | 0.2973 | 0.1973 | 1.32E-01 | 1.00E+00 |
| Other immunodeficiency disorder | 1.81 | (0.53, 6.16) | 0.5907 | 0.3788 | 1.19E-01 | 1.00E+00 |
| **Other lung condition** | 3.29 | (1.35, 8.00) | 1.1909 | 0.274 | 1.39E-05 | 5.82E-04 |
| **Any** | 2.85 | (1.78, 4.57) | 1.0485 | 0.1452 | 5.22E-13 | 2.19E-11 |
| **OBEESITY** |  |  |  |  |  |  |
| **obesity I (BMI >= 30 and BMI < 35)** | 1.31 | (0.76, 2.27) | 0.2714 | 0.1696 | 1.09E-01 | 1.00E+00 |
| **obesity II (BMI >= 35 and BMI < 40)** | 1.16 | (0.56, 2.43) | 0.1525 | 0.2275 | 5.03E-01 | 1.00E+00 |
| **obesity III (BMI >= 40)** | 2.06 | (1.05, 4.02) | 0.7211 | 0.2071 | 4.98E-04 | 2.09E-02 |
| **Not obese** | 1 |  | 0 |  | nan | nan |
| **GENETIC SEX** |  |  |  |  |  |  |
| **Female** | 1 |  | 0 |  | nan | nan |
| **Male** | 1.54 | (1.00, 2.37) | 0.4342 | 0.1325 | 1.05E-03 | 4.42E-02 |
| **SYMPTOMS** |  |  |  |  |  |  |
| **Abdominal pain** | 1.95 | (1.23, 3.08) | 0.666 | 0.1413 | 2.45E-06 | 1.03E-04 |
| **Body aches** | 1.58 | (0.95, 2.60) | 0.4547 | 0.1546 | 3.27E-03 | 1.37E-01 |
| **Change in taste or smell** | 0.79 | (0.51, 1.23) | -0.2319 | 0.1358 | 8.79E-02 | 1.00E+00 |
| **Chills** | 2.04 | (1.28, 3.23) | 0.7106 | 0.1423 | 5.96E-07 | 2.50E-05 |
| **Cough producing phlegm** | 1.83 | (1.18, 2.84) | 0.6063 | 0.1351 | 7.24E-06 | 3.04E-04 |
| **Diarrhea** | 2.48 | (1.61, 3.84) | 0.9102 | 0.1339 | 1.08E-11 | 4.53E-10 |
| **Dry cough** | 2.67 | (1.66, 4.31) | 0.9836 | 0.1474 | 2.49E-11 | 1.04E-09 |
| **Feeling tired or fatigue** | 2.55 | (1.09, 5.97) | 0.9344 | 0.2628 | 3.77E-04 | 1.58E-02 |
| **Fever** | 3.52 | (2.08, 5.96) | 1.2595 | 0.1622 | 8.00E-15 | 3.36E-13 |
| **Headache** | 0.97 | (0.61, 1.52) | -0.0356 | 0.1402 | 8.00E-01 | 1.00E+00 |
| **Nasal congestion** | 1.06 | (0.69, 1.64) | 0.0592 | 0.1337 | 6.58E-01 | 1.00E+00 |
| **Nausea or vomiting** | 2.27 | (1.44, 3.60) | 0.8217 | 0.1415 | 6.30E-09 | 2.64E-07 |
| **Runny nose** | 1.31 | (0.83, 2.07) | 0.2696 | 0.1415 | 5.68E-02 | 1.00E+00 |
| **Shortness of breath** | 11.55 | (5.91, 22.59) | 2.4466 | 0.2069 | 2.93E-32 | 1.23E-30 |
| **Sore throat** | 1.45 | (0.92, 2.29) | 0.374 | 0.1396 | 7.40E-03 | 3.11E-01 |

Unadjusted odds ratios for progression of COVID-19 infection to a critical, life-threatening illness were calculated among those reporting a positive COVID-19 test result. The following groups served as reference populations for the odds ratio calculations: European genetic ancestry (Genetic Ancestry), Ages 50-64 (Age), Female (Genetic Sex), and Not Obese (BMI < 30, Obesity). Note that symptoms odds ratios were calculated in a binary fashion among symptomatic customers only (Methods).

**Supplementary Table 14.** Risk factor odds ratios for progression of COVID-19 to a critical case, adjusted for health conditions, obesity, age, and sex.

| RISK FACTOR | ODDS RATIO (OR) | OR 95% CI (BONFERRONI) | logOR | logOR S.E. | P-VALUE (NOMINAL) | P-VALUE (BONFERRONI) |
| --- | --- | --- | --- | --- | --- | --- |
| **AGE** |  |  |  |  |  |  |
| Ages 18-29 | 0.46 | (0.15, 1.45) | -0.7675 | 0.3506 | 2.86E-02 | 1.00E+00 |
| Ages 30-49 | 0.67 | (0.38, 1.17) | -0.4033 | 0.1735 | 2.01E-02 | 8.45E-01 |
| Ages 50-64 | 1 |  | 0 |  | nan | nan |
| Ages 65+ | 1.51 | (0.85, 2.69) | 0.41 | 0.1785 | 2.16E-02 | 9.07E-01 |
| **GENETIC ANCESTRY** |  |  |  |  |  |  |
| Admixed African-European | 2.34 | (1.11, 4.95) | 0.8516 | 0.2308 | 2.25E-04 | 9.43E-03 |
| European | 1 |  | 0 |  | nan | nan |
| Admixed Amerindian | 1.12 | (0.49, 2.56) | 0.1145 | 0.2548 | 6.53E-01 | 1.00E+00 |
| Other | 0.85 | (0.44, 1.65) | -0.164 | 0.206 | 4.26E-01 | 1.00E+00 |
| **EXPOSURE** |  |  |  |  |  |  |
| Any | 0.83 | (0.41, 1.67) | -0.1862 | 0.2163 | 3.89E-01 | 1.00E+00 |
| Biological relative tested positive | 1.23 | (0.73, 2.08) | 0.2104 | 0.1609 | 1.91E-01 | 1.00E+00 |
| Directly exposed to someone who tested positive | 0.62 | (0.34, 1.13) | -0.4747 | 0.183 | 9.49E-03 | 3.98E-01 |
| Healthcare worker directly exposed | 0.62 | (0.31, 1.24) | -0.4703 | 0.2116 | 2.63E-02 | 1.00E+00 |
| Household member tested positive | 1.01 | (0.63, 1.60) | 0.0068 | 0.1434 | 9.62E-01 | 1.00E+00 |
| **HEALTH CONDITIONS** |  |  |  |  |  |  |
| Asthma | 1.24 | (0.69, 2.22) | 0.2124 | 0.1806 | 2.40E-01 | 1.00E+00 |
| COPD (Chronic Obstructive Pulmonary Disease) | 2.39 | (0.90, 6.36) | 0.8704 | 0.302 | 3.95E-03 | 1.66E-01 |
| Cancer treated in the past year | 1.08 | (0.26, 4.55) | 0.0784 | 0.4429 | 8.59E-01 | 1.00E+00 |
| Cardiovascular disease | 1.42 | (0.64, 3.15) | 0.3485 | 0.2469 | 1.58E-01 | 1.00E+00 |
| Chronic kidney disease | 4.88 | (1.67, 14.30) | 1.586 | 0.3314 | 1.70E-06 | 7.13E-05 |
| Diabetes | 2.16 | (1.20, 3.89) | 0.7699 | 0.1816 | 2.23E-05 | 9.38E-04 |
| Hypertension | 1.34 | (0.80, 2.24) | 0.2903 | 0.1599 | 6.95E-02 | 1.00E+00 |
| Other | 1.19 | (0.49, 2.88) | 0.1765 | 0.2714 | 5.16E-01 | 1.00E+00 |
| Other autoimmune disease | 1.55 | (0.78, 3.06) | 0.4368 | 0.2105 | 3.80E-02 | 1.00E+00 |
| Other immunodeficiency disorder | 1.82 | (0.52, 6.31) | 0.5962 | 0.3844 | 1.21E-01 | 1.00E+00 |
| **Other lung condition** | 2.09 | (0.80, 5.47) | 0.7376 | 0.2965 | 1.29E-02 | 5.40E-01 |
| **Any** | 2.22 | (1.33, 3.69) | 0.7959 | 0.157 | 4.00E-07 | 1.68E-05 |
| **OBESITY** |  |  |  |  |  |  |
| **obesity I (BMI ≥ 30 and BMI < 35)** | 1.09 | (0.62, 1.93) | 0.0882 | 0.1761 | 6.17E-01 | 1.00E+00 |
| **obesity II (BMI ≥ 35 and BMI < 40)** | 1.06 | (0.50, 2.26) | 0.0605 | 0.2334 | 7.96E-01 | 1.00E+00 |
| **obesity III (BMI ≥ 40)** | 1.95 | (0.96, 3.98) | 0.6695 | 0.2196 | 2.29E-03 | 9.64E-02 |
| **Not obese** | 1 |  | 0 |  | nan | nan |
| **GENETIC SEX** |  |  |  |  |  |  |
| **Female** | 1 |  | 0 |  | nan | nan |
| **Male** | 1.5 | (0.95, 2.37) | 0.4041 | 0.142 | 4.42E-03 | 1.85E-01 |
| **SYMPTOMS** |  |  |  |  |  |  |
| **Abdominal pain** | 2.03 | (1.24, 3.32) | 0.7095 | 0.1514 | 2.77E-06 | 1.16E-04 |
| **Body aches** | 1.88 | (1.08, 3.27) | 0.6321 | 0.1707 | 2.13E-04 | 8.95E-03 |
| **Change in taste or smell** | 1.03 | (0.63, 1.66) | 0.025 | 0.1485 | 8.66E-01 | 1.00E+00 |
| **Chills** | 2.22 | (1.35, 3.66) | 0.7975 | 0.1542 | 2.31E-07 | 9.70E-06 |
| **Cough producing phlegm** | 1.69 | (1.05, 2.70) | 0.5233 | 0.1452 | 3.14E-04 | 1.32E-02 |
| **Diarrhea** | 2.74 | (1.71, 4.39) | 1.0077 | 0.1457 | 4.58E-12 | 1.92E-10 |
| **Dry cough** | 2.73 | (1.62, 4.59) | 1.0041 | 0.1601 | 3.60E-10 | 1.51E-08 |
| **Feeling tired or fatigue** | 2.86 | (1.10, 7.40) | 1.05 | 0.2938 | 3.51E-04 | 1.48E-02 |
| **Fever** | 3.34 | (1.91, 5.85) | 1.2074 | 0.1726 | 2.65E-12 | 1.11E-10 |
| **Headache** | 1.18 | (0.72, 1.95) | 0.1678 | 0.1537 | 2.75E-01 | 1.00E+00 |
| **Nasal congestion** | 1.12 | (0.70, 1.78) | 0.1106 | 0.1435 | 4.41E-01 | 1.00E+00 |
| **Nausea or vomiting** | 2.72 | (1.65, 4.49) | 0.9997 | 0.1547 | 1.03E-10 | 4.31E-09 |
| **Runny nose** | 1.19 | (0.72, 1.95) | 0.1704 | 0.153 | 2.65E-01 | 1.00E+00 |
| **Shortness of breath** | 12.87 | (6.18, 26.81) | 2.5549 | 0.2265 | 1.62E-29 | 6.79E-28 |
| **Sore throat** | 1.65 | (1.01, 2.68) | 0.4991 | 0.1503 | 9.01E-04 | 3.79E-02 |

Adjusted odds ratios for progression of COVID-19 infection to a critical, life-threatening illness were calculated among those reporting a positive COVID-19 test result. The following groups served as reference populations for the odds ratio calculations: European genetic ancestry (Genetic Ancestry), Ages 50-64 (Age), Female (Genetic Sex), and Not Obese (BMI < 30, Obesity). Note that symptoms odds ratios were calculated in a binary fashion among symptomatic customers only (Methods). Here, odds ratios were adjusted for age, sex, comorbidities (Y/N if any), and obesity (Y/N if BMI > 30, Methods).

**Supplementary Table 15.** Unadjusted risk factor odds ratios for COVID-19 susceptibility (training dataset)

| RISK FACTOR | ODDS RATIO (OR) | OR 95% CI (BONFERRONI) | logOR | logOR S.E. | P-VALUE (NOMINAL) | P-VALUE (BONFERRONI) |
| --- | --- | --- | --- | --- | --- | --- |
| **AGE** |  |  |  |  |  |  |
| Ages 18-29 | 1.6 | (1.21, 2.11) | 0.4693 | 0.0857 | 4.42E-08 | 1.86E-06 |
| Ages 30-49 | 1.11 | (0.92, 1.34) | 0.105 | 0.0584 | 7.22E-02 | 1.00E+00 |
| Ages 50-64 | 1 |  | 0 |  | nan | nan |
| Ages 65+ | 0.49 | (0.38, 0.63) | -0.7051 | 0.077 | 0.00E+00 | 0.00E+00 |
| **GENETIC ANCESTRY** |  |  |  |  |  |  |
| Admixed African-European | 1.24 | (0.85, 1.81) | 0.2158 | 0.1168 | 6.47E-02 | 1.00E+00 |
| European | 1 |  | 0 |  | nan | nan |
| Admixed Amerindian | 1.59 | (1.22, 2.07) | 0.462 | 0.0814 | 1.36E-08 | 5.73E-07 |
| Other | 1.16 | (0.94, 1.44) | 0.1516 | 0.0665 | 2.27E-02 | 9.52E-01 |
| **EXPOSURE** |  |  |  |  |  |  |
| Any | 9.49 | (7.65, 11.77) | 2.2503 | 0.0665 | 4.79E-251 | 2.01E-249 |
| Biological relative tested positive | 5.74 | (4.66, 7.07) | 1.7467 | 0.0644 | 4.71E-162 | 1.98E-160 |
| Directly exposed to someone who tested positive | 7.63 | (6.33, 9.19) | 2.0317 | 0.0576 | 4.44E-272 | 1.87E-270 |
| Healthcare worker directly exposed | 1.56 | (1.29, 1.87) | 0.4417 | 0.0576 | 1.78E-14 | 7.47E-13 |
| Household member tested positive | 24.28 | (19.62, 30.03) | 3.1896 | 0.0657 | 0.00E+00 | 0.00E+00 |
| **PRE-EXISTING CONDITION** |  |  |  |  |  |  |
| Asthma | 0.81 | (0.65, 1.00) | -0.2166 | 0.0677 | 1.37E-03 | 5.75E-02 |
| COPD (Chronic Obstructive Pulmonary Disease) | 0.54 | (0.31, 0.93) | -0.6217 | 0.1684 | 2.22E-04 | 9.32E-03 |
| Cancer treated in the past year | 0.57 | (0.32, 1.00) | -0.564 | 0.1731 | 1.12E-03 | 4.71E-02 |
| Cardiovascular disease | 0.74 | (0.51, 1.06) | -0.3014 | 0.1122 | 7.26E-03 | 3.05E-01 |
| Chronic kidney disease | 0.55 | (0.28, 1.10) | -0.5938 | 0.2139 | 5.49E-03 | 2.31E-01 |
| Diabetes | 0.9 | (0.69, 1.17) | -0.105 | 0.0816 | 1.98E-01 | 1.00E+00 |
| Hypertension | 0.76 | (0.62, 0.93) | -0.2728 | 0.061 | 7.64E-06 | 3.21E-04 |
| Other | 0.93 | (0.66, 1.31) | -0.0726 | 0.1049 | 4.89E-01 | 1.00E+00 |
| Other autoimmune disease | 0.8 | (0.61, 1.04) | -0.2241 | 0.081 | 5.65E-03 | 2.37E-01 |
| Other immunodeficiency disorder | 0.77 | (0.42, 1.40) | -0.2669 | 0.1854 | 1.50E-01 | 1.00E+00 |
| Other lung condition | 0.64 | (0.39, 1.03) | -0.4524 | 0.1502 | 2.59E-03 | 1.09E-01 |
| Any | 0.67 | (0.57, 0.79) | -0.3983 | 0.0498 | 1.33E-15 | 5.60E-14 |
| **OBESITY** |  |  |  |  |  |  |
| obesity I (BMI >= 30 and BMI < 35) | 1.02 | (0.83, 1.25) | 0.0212 | 0.0628 | 7.35E-01 | 1.00E+00 |
| obesity II (BMI >= 35 and BMI < 40) | 1.15 | (0.94, 1.41) | 0.1376 | 0.0626 | 2.81E-02 | 1.00E+00 |
| obesity III (BMI >= 40) | 1.13 | (0.85, 1.50) | 0.12 | 0.0874 | 1.70E-01 | 1.00E+00 |
| Not obese | 1 |  | 0 |  | nan | nan |
| **GENETIC SEX** |  |  |  |  |  |  |
| Female | 1 |  | 0 |  | nan | nan |
| Male | 1.16 | (0.98, 1.37) | 0.1459 | 0.0525 | 5.43E-03 | 2.28E-01 |
| **SYMPTOMS** |  |  |  |  |  |  |
| Abdominal pain | 0.97 | (0.64, 1.48) | -0.0303 | 0.1303 | 8.16E-01 | 1.00E+00 |
| Body aches | 1.56 | (1.09, 2.23) | 0.4461 | 0.1104 | 5.34E-05 | 2.24E-03 |
| Change in taste or smell | 7.02 | (4.67, 10.56) | 1.9493 | 0.1257 | 2.96E-54 | 1.24E-52 |
| Chills | 1.38 | (0.97, 1.96) | 0.3211 | 0.1085 | 3.08E-03 | 1.29E-01 |
| Cough producing phlegm | 0.93 | (0.64, 1.37) | -0.0685 | 0.1178 | 5.61E-01 | 1.00E+00 |
| Diarrhea | 1.18 | (0.81, 1.72) | 0.1672 | 0.1158 | 1.49E-01 | 1.00E+00 |
| Dry cough | 1.29 | (0.91, 1.84) | 0.2578 | 0.1082 | 1.72E-02 | 7.21E-01 |
| Feeling tired or fatigue | 1.54 | (0.99, 2.42) | 0.4341 | 0.1381 | 1.67E-03 | 7.03E-02 |
| Fever | 1.54 | (1.08, 2.19) | 0.4295 | 0.1088 | 7.84E-05 | 3.29E-03 |
| Headache | 1.39 | (0.97, 2.00) | 0.33 | 0.1118 | 3.17E-03 | 1.33E-01 |
| Nasal congestion | 0.85 | (0.60, 1.21) | -0.1632 | 0.1081 | 1.31E-01 | 1.00E+00 |
| Nausea or vomiting | 0.82 | (0.53, 1.26) | -0.1979 | 0.1327 | 1.36E-01 | 1.00E+00 |
| Runny nose | 0.59 | (0.41, 0.86) | -0.5246 | 0.1137 | 3.92E-06 | 1.65E-04 |
| Shortness of breath | 1.25 | (0.87, 1.79) | 0.2219 | 0.1106 | 4.48E-02 | 1.00E+00 |
| Sore throat | 0.48 | (0.34, 0.69) | -0.7286 | 0.1117 | 6.84E-11 | 2.87E-09 |

Unadjusted odds ratios for a positive COVID-19 test were calculated among survey participants reporting a test result (positive or negative). The following groups served as reference populations for the odds ratio calculations: European genetic ancestry (Genetic Ancestry), Ages 50-64 (Age), Female (Genetic Sex), and Not Obese (BMI < 30, Obesity). Note that symptoms odds ratios were calculated in a binary fashion among symptomatic customers only (Methods).

**Supplementary Table 16.** Unadjusted risk factor odds ratios for COVID-19 hospitalization (training dataset)

| RISK FACTOR | ODDS RATIO (OR) | OR 95% CI (BONFERRONI) | logOR | logOR S.E. | P-VALUE (NOMINAL) | P-VALUE (BONFERRONI) |
| --- | --- | --- | --- | --- | --- | --- |
| **AGE** |  |  |  |  |  |  |
| Ages 18-29 | 0.53 | (0.20, 1.37) | -0.6395 | 0.2951 | 3.02E-02 | 1.00E+00 |
| Ages 30-49 | 0.5 | (0.27, 0.92) | -0.6907 | 0.1879 | 2.37E-04 | 9.95E-03 |
| Ages 50-64 | 1 |  | 0 |  | nan | nan |
| Ages 65+ | 1.64 | (0.87, 3.08) | 0.4956 | 0.1943 | 1.07E-02 | 4.51E-01 |
| **GENETIC ANCESTRY** |  |  |  |  |  |  |
| Admixed African-European | 1.19 | (0.43, 3.28) | 0.176 | 0.3121 | 5.73E-01 | 1.00E+00 |
| European | 1 |  | 0 |  | nan | nan |
| Admixed Amerindian | 0.77 | (0.32, 1.86) | -0.2551 | 0.2703 | 3.45E-01 | 1.00E+00 |
| Other | 0.69 | (0.34, 1.39) | -0.3752 | 0.2169 | 8.37E-02 | 1.00E+00 |
| **EXPOSURE** |  |  |  |  |  |  |
| Any | 0.69 | (0.34, 1.39) | -0.3732 | 0.2174 | 8.61E-02 | 1.00E+00 |
| Biological relative tested positive | 1.06 | (0.60, 1.88) | 0.0572 | 0.1772 | 7.47E-01 | 1.00E+00 |
| Directly exposed to someone who tested positive | 0.54 | (0.29, 0.98) | -0.6207 | 0.1867 | 8.86E-04 | 3.72E-02 |
| Healthcare worker directly exposed | 0.57 | (0.29, 1.10) | -0.5698 | 0.2042 | 5.26E-03 | 2.21E-01 |
| Household member tested positive | 0.88 | (0.53, 1.45) | -0.1332 | 0.1551 | 3.90E-01 | 1.00E+00 |
| **HEALTH CONDITIONS** |  |  |  |  |  |  |
| Asthma | 1.17 | (0.62, 2.19) | 0.1555 | 0.1944 | 4.24E-01 | 1.00E+00 |
| COPD (Chronic Obstructive Pulmonary Disease) | 2.95 | (0.82, 10.67) | 1.083 | 0.3962 | 6.27E-03 | 2.63E-01 |
| Cancer treated in the past year | 3.21 | (0.88, 11.78) | 1.1677 | 0.4006 | 3.56E-03 | 1.49E-01 |
| Cardiovascular disease | 2.73 | (1.18, 6.28) | 1.0036 | 0.2573 | 9.62E-05 | 4.04E-03 |
| Chronic kidney disease | 10.82 | (2.86, 40.90) | 2.3812 | 0.4103 | 6.49E-09 | 2.73E-07 |
| Diabetes | 3.21 | (1.70, 6.06) | 1.1676 | 0.1957 | 2.44E-09 | 1.02E-07 |
| Hypertension | 1.68 | (0.98, 2.88) | 0.5172 | 0.1664 | 1.88E-03 | 7.91E-02 |
| Other | 1.63 | (0.74, 3.63) | 0.4906 | 0.2463 | 4.64E-02 | 1.00E+00 |
| Other autoimmune disease | 1.21 | (0.58, 2.54) | 0.1914 | 0.2288 | 4.03E-01 | 1.00E+00 |
| Other immunodeficiency disorder | 0.77 | (0.11, 5.53) | -0.2655 | 0.6094 | 6.63E-01 | 1.00E+00 |
| **Other lung condition** | 1.8 | (0.50, 6.55) | 0.5883 | 0.3983 | 1.40E-01 | 1.00E+00 |
| **Any** | 2.36 | (1.41, 3.93) | 0.8574 | 0.1575 | 5.26E-08 | 2.21E-06 |
| **OBESITY** |  |  |  |  |  |  |
| **obesity I (BMI ≥ 30 and BMI < 35)** | 1.32 | (0.74, 2.34) | 0.2765 | 0.1771 | 1.19E-01 | 1.00E+00 |
| **obesity II (BMI ≥ 35 and BMI < 40)** | 1.08 | (0.56, 2.06) | 0.0753 | 0.2001 | 7.07E-01 | 1.00E+00 |
| **obesity III (BMI ≥ 40)** | 1.42 | (0.62, 3.26) | 0.3505 | 0.2566 | 1.72E-01 | 1.00E+00 |
| **Not obese** | 1 |  | 0 |  | nan | nan |
| **GENETIC SEX** |  |  |  |  |  |  |
| **Female** | 1 |  | 0 |  | nan | nan |
| **Male** | 1.29 | (0.79, 2.12) | 0.2584 | 0.1525 | 9.02E-02 | 1.00E+00 |
| **SYMPTOMS** |  |  |  |  |  |  |
| **Abdominal pain** | 1.79 | (1.04, 3.10) | 0.5841 | 0.1686 | 5.33E-04 | 2.24E-02 |
| **Body aches** | 1.6 | (0.90, 2.83) | 0.4695 | 0.1765 | 7.80E-03 | 3.28E-01 |
| **Change in taste or smell** | 0.83 | (0.50, 1.38) | -0.189 | 0.1582 | 2.32E-01 | 1.00E+00 |
| **Chills** | 2.18 | (1.29, 3.71) | 0.7814 | 0.1632 | 1.68E-06 | 7.07E-05 |
| **Cough producing phlegm** | 1.51 | (0.90, 2.53) | 0.4096 | 0.1596 | 1.03E-02 | 4.31E-01 |
| **Diarrhea** | 2.04 | (1.24, 3.36) | 0.7114 | 0.154 | 3.87E-06 | 1.62E-04 |
| **Dry cough** | 2.5 | (1.47, 4.28) | 0.9179 | 0.1653 | 2.80E-08 | 1.18E-06 |
| **Feeling tired or fatigue** | 4.47 | (1.35, 14.73) | 1.4965 | 0.3681 | 4.79E-05 | 2.01E-03 |
| **Fever** | 3.06 | (1.72, 5.44) | 1.1178 | 0.178 | 3.36E-10 | 1.41E-08 |
| **Headache** | 0.87 | (0.52, 1.47) | -0.1351 | 0.16 | 3.99E-01 | 1.00E+00 |
| **Nasal congestion** | 0.73 | (0.44, 1.23) | -0.3139 | 0.1595 | 4.90E-02 | 1.00E+00 |
| **Nausea or vomiting** | 2.48 | (1.45, 4.23) | 0.9068 | 0.1653 | 4.13E-08 | 1.74E-06 |
| **Runny nose** | 0.87 | (0.49, 1.55) | -0.1355 | 0.1777 | 4.46E-01 | 1.00E+00 |
| **Shortness of breath** | 6.4 | (3.49, 11.73) | 1.8558 | 0.187 | 3.34E-23 | 1.40E-21 |
| **Sore throat** | 1.03 | (0.59, 1.80) | 0.0332 | 0.1717 | 8.47E-01 | 1.00E+00 |

Unadjusted odds ratios for hospitalization due to COVID-19 infection were calculated among those reporting a positive COVID-19 test result. The following groups served as reference populations for the odds ratio calculations: European genetic ancestry (Genetic Ancestry), Ages 50-64 (Age), Female (Genetic Sex), and Not Obese (BMI < 30, Obesity). Note that symptoms odds ratios were calculated in a binary fashion among symptomatic customers only (Methods).

**Supplementary Table 17.**
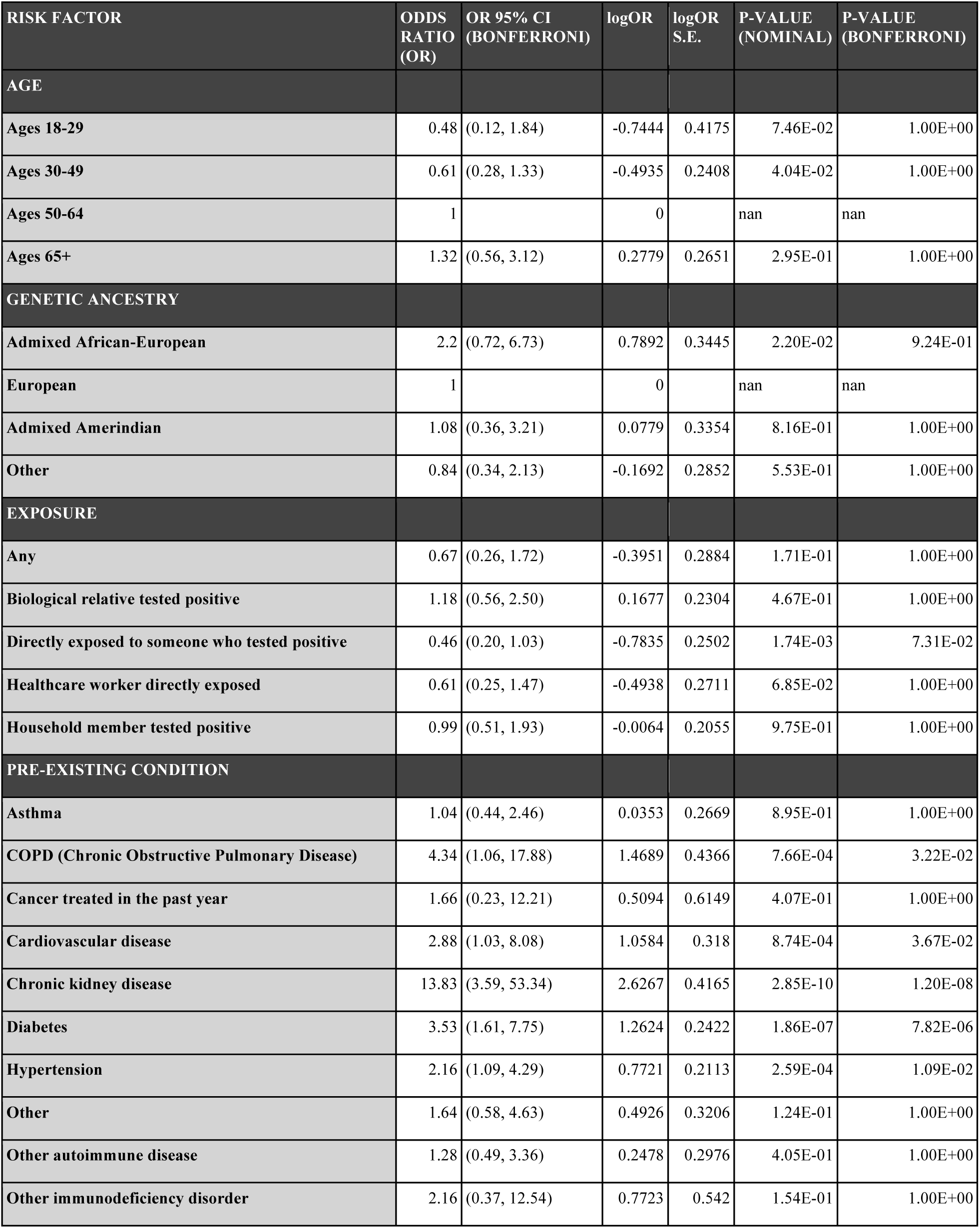
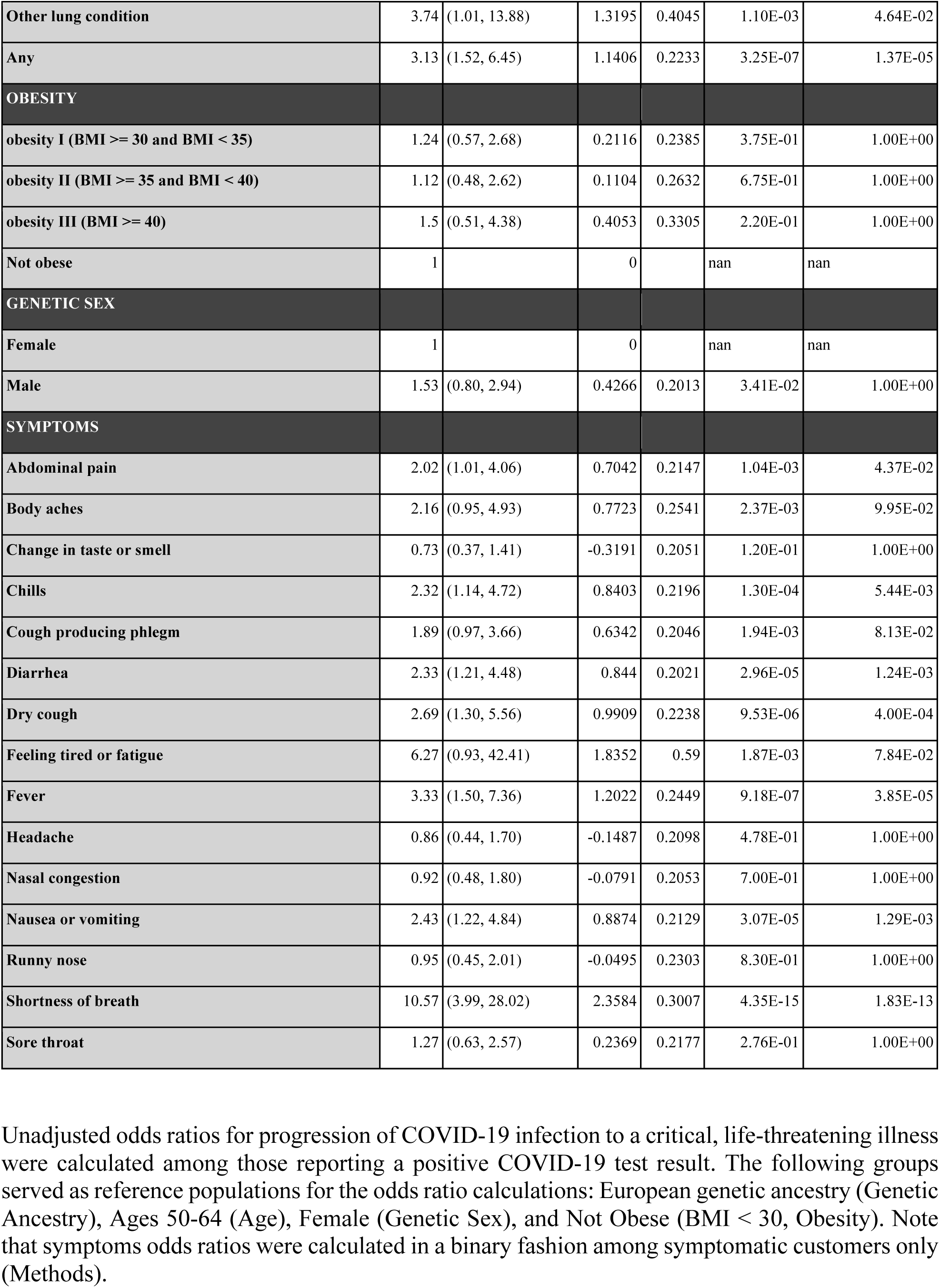
Unadjusted risk factor odds ratios for progression of COVID-19 to a critical case (training dataset)

**Supplementary Table 18.**
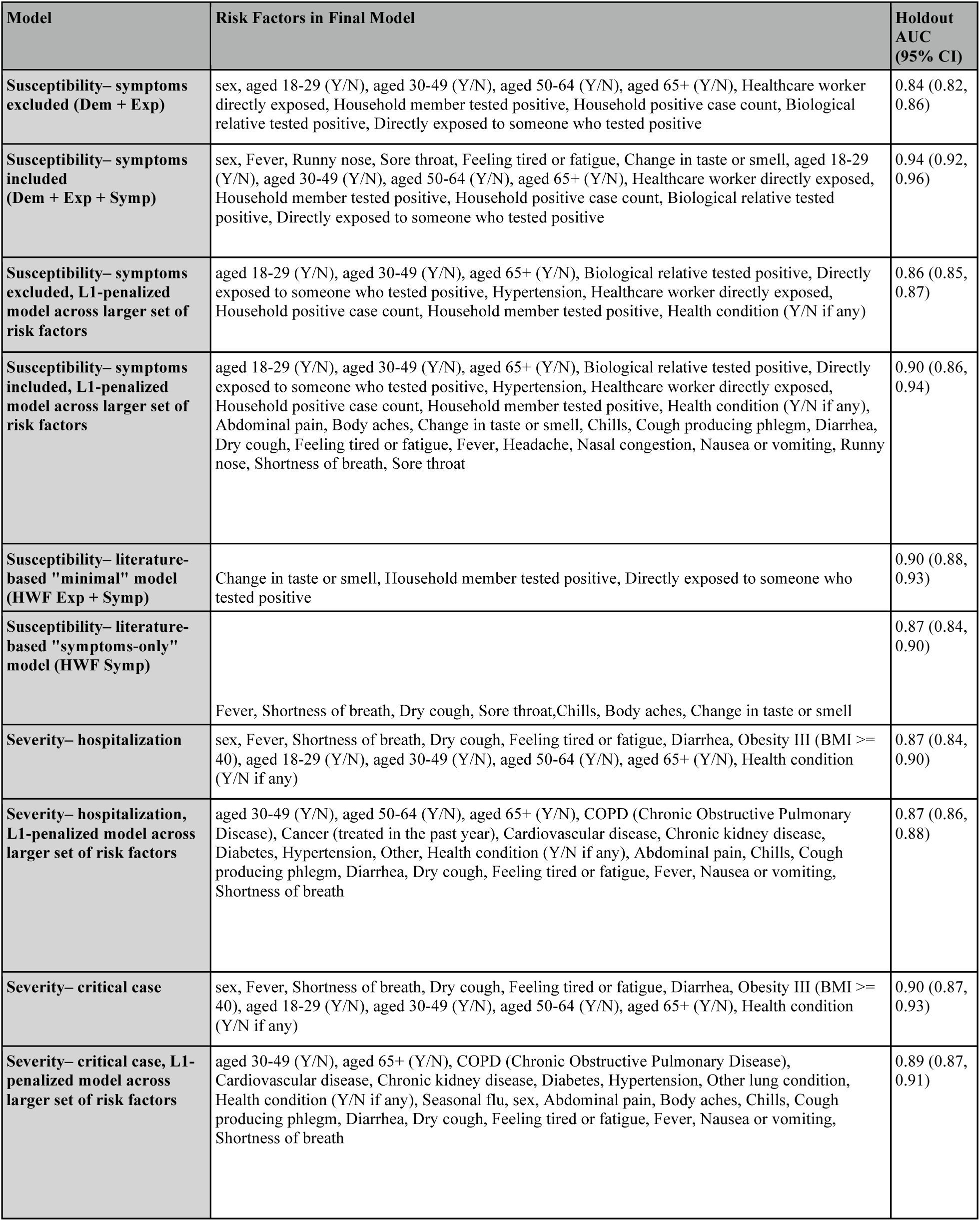
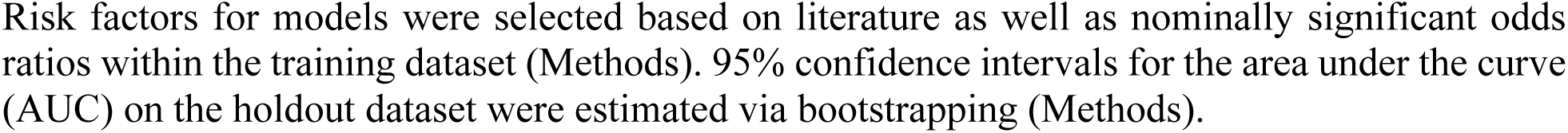
Summary of risk factors and performances of different models.

| Model | Risk Factors in Final Model | Holdout AUC (95% CI) |
| --- | --- | --- |
| **Susceptibility– symptoms excluded (Dem + Exp)** | sex, aged 18-29 (Y/N), aged 30-49 (Y/N), aged 50-64 (Y/N), aged 65+ (Y/N), Healthcare worker directly exposed, Household member tested positive, Household positive case count, Biological relative tested positive, Directly exposed to someone who tested positive | 0.84 (0.82, 0.86) |
| **Susceptibility– symptoms included (Dem + Exp + Symp)** | sex, Fever, Runny nose, Sore throat, Feeling tired or fatigue, Change in taste or smell, aged 18-29 (Y/N), aged 30-49 (Y/N), aged 50-64 (Y/N), aged 65+ (Y/N), Healthcare worker directly exposed, Household member tested positive, Household positive case count, Biological relative tested positive, Directly exposed to someone who tested positive | 0.94 (0.92, 0.96) |
| **Susceptibility– symptoms excluded, L1-penalized model across larger set of risk factors** | aged 18-29 (Y/N), aged 30-49 (Y/N), aged 65+ (Y/N), Biological relative tested positive, Directly exposed to someone who tested positive, Hypertension, Healthcare worker directly exposed, Household positive case count, Household member tested positive, Health condition (Y/N if any) | 0.86 (0.85, 0.87) |
| **Susceptibility– symptoms included, L1-penalized model across larger set of risk factors** | aged 18-29 (Y/N), aged 30-49 (Y/N), aged 65+ (Y/N), Biological relative tested positive, Directly exposed to someone who tested positive, Hypertension, Healthcare worker directly exposed, Household positive case count, Household member tested positive, Health condition (Y/N if any), Abdominal pain, Body aches, Change in taste or smell, Chills, Cough producing phlegm, Diarrhea, Dry cough, Feeling tired or fatigue, Fever, Headache, Nasal congestion, Nausea or vomiting, Runny nose, Shortness of breath, Sore throat | 0.90 (0.86, 0.94) |
| **Susceptibility– literature-based "minimal" model (HWF Exp + Symp)** | Change in taste or smell, Household member tested positive, Directly exposed to someone who tested positive | 0.90 (0.88, 0.93) |
| **Susceptibility– literature-based "symptoms-only" model (HWF Symp)** | Fever, Shortness of breath, Dry cough, Sore throat, Chills, Body aches, Change in taste or smell | 0.87 (0.84, 0.90) |
| **Severity– hospitalization** | sex, Fever, Shortness of breath, Dry cough, Feeling tired or fatigue, Diarrhea, Obesity III (BMI ≥ 40), aged 18-29 (Y/N), aged 30-49 (Y/N), aged 50-64 (Y/N), aged 65+ (Y/N), Health condition (Y/N if any) | 0.87 (0.84, 0.90) |
| **Severity– hospitalization, L1-penalized model across larger set of risk factors** | aged 30-49 (Y/N), aged 50-64 (Y/N), aged 65+ (Y/N), COPD (Chronic Obstructive Pulmonary Disease), Cancer (treated in the past year), Cardiovascular disease, Chronic kidney disease, Diabetes, Hypertension, Other, Health condition (Y/N if any), Abdominal pain, Chills, Cough producing phlegm, Diarrhea, Dry cough, Feeling tired or fatigue, Fever, Nausea or vomiting, Shortness of breath | 0.87 (0.86, 0.88) |
| **Severity– critical case** | sex, Fever, Shortness of breath, Dry cough, Feeling tired or fatigue, Diarrhea, Obesity III (BMI ≥ 40), aged 18-29 (Y/N), aged 30-49 (Y/N), aged 50-64 (Y/N), aged 65+ (Y/N), Health condition (Y/N if any) | 0.90 (0.87, 0.93) |
| **Severity– critical case, L1-penalized model across larger set of risk factors** | aged 30-49 (Y/N), aged 65+ (Y/N), COPD (Chronic Obstructive Pulmonary Disease), Cardiovascular disease, Chronic kidney disease, Diabetes, Hypertension, Other lung condition, Health condition (Y/N if any), Seasonal flu, sex, Abdominal pain, Body aches, Chills, Cough producing phlegm, Diarrhea, Dry cough, Feeling tired or fatigue, Fever, Nausea or vomiting, Shortness of breath | 0.89 (0.87, 0.91) |

Risk factors for models were selected based on literature as well as nominally significant odds ratios within the training dataset (Methods). 95% confidence intervals for the area under the curve (AUC) on the holdout dataset were estimated via bootstrapping (Methods).

**Supplementary Table 19.** Stratified holdout performance assessment for a COVID-19 susceptibility model built from age, sex, and exposures (Dem + Exp)

| cohort | N | ncase | ncontrol | AUC | Sensitivity | Specificity |
| --- | --- | --- | --- | --- | --- | --- |
| Admixed African-European Genetic Ancestry | 228 | 38 | 190 | 0.88 (0.82, 0.94) | 0.89 (0.80, 0.99) | 0.65 (0.59, 0.72) |
| All | 4834 | 632 | 4202 | 0.84 (0.82, 0.86) | 0.75 (0.72, 0.79) | 0.76 (0.75, 0.78) |
| European Genetic Ancestry | 3372 | 408 | 2964 | 0.84 (0.82, 0.87) | 0.74 (0.70, 0.78) | 0.79 (0.77, 0.80) |
| Admixed Amerindian Genetic Ancestry | 410 | 73 | 337 | 0.80 (0.74, 0.86) | 0.74 (0.64, 0.84) | 0.71 (0.66, 0.75) |
| Other Genetic Ancestry | 824 | 113 | 711 | 0.83 (0.78, 0.87) | 0.77 (0.69, 0.85) | 0.71 (0.68, 0.75) |
| Ages 18-29 | 340 | 74 | 266 | 0.83 (0.77, 0.89) | 0.81 (0.72, 0.90) | 0.65 (0.59, 0.71) |
| Ages 30-49 | 1744 | 270 | 1474 | 0.80 (0.77, 0.83) | 0.82 (0.77, 0.86) | 0.59 (0.56, 0.61) |
| Ages 50-65 | 1515 | 200 | 1315 | 0.85 (0.82, 0.88) | 0.73 (0.67, 0.79) | 0.81 (0.79, 0.83) |
| Ages 65+ | 1235 | 88 | 1147 | 0.81 (0.75, 0.87) | 0.57 (0.47, 0.67) | 0.96 (0.95, 0.97) |
| Female | 3354 | 409 | 2945 | 0.83 (0.81, 0.85) | 0.73 (0.68, 0.77) | 0.77 (0.75, 0.79) |
| Male | 1480 | 223 | 1257 | 0.85 (0.82, 0.88) | 0.81 (0.76, 0.86) | 0.75 (0.73, 0.77) |

**Supplementary Table 20.** Stratified holdout performance assessment for a COVID-19 susceptibility model built from age, sex, exposures, and symptoms (Dem + Exp + Symp)

| cohort | N | ncase | ncontrol | AUC | Sensitivity | Specificity |
| --- | --- | --- | --- | --- | --- | --- |
| Admixed African-European Genetic Ancestry | 96 | 12 | 84 | 0.96 (0.88, 1.00) | 0.91 (0.75, 1.00) | 0.92 (0.86, 0.98) |
| All | 2047 | 229 | 1818 | 0.94 (0.92, 0.96) | 0.85 (0.80, 0.89) | 0.91 (0.90, 0.93) |
| European Genetic Ancestry | 1457 | 146 | 1311 | 0.93 (0.90, 0.96) | 0.83 (0.77, 0.90) | 0.91 (0.90, 0.93) |
| Admixed Amerindian Genetic Ancestry | 155 | 23 | 132 | 0.98 (0.95, 1.00) | 0.96 (0.87, 1.00) | 0.89 (0.84, 0.95) |
| Other Genetic Ancestry | 339 | 48 | 291 | 0.94 (0.90, 0.99) | 0.81 (0.70, 0.92) | 0.93 (0.90, 0.96) |
| Ages 18-29 | 132 | 29 | 103 | 0.98 (0.95, 1.00) | 0.93 (0.84, 1.00) | 0.92 (0.87, 0.97) |
| Ages 30-49 | 582 | 77 | 505 | 0.96 (0.95, 0.98) | 0.89 (0.82, 0.96) | 0.88 (0.85, 0.91) |
| Ages 50-65 | 616 | 83 | 533 | 0.93 (0.90, 0.97) | 0.84 (0.76, 0.92) | 0.91 (0.88, 0.93) |
| Ages 65+ | 717 | 40 | 677 | 0.89 (0.83, 0.96) | 0.72 (0.58, 0.86) | 0.95 (0.93, 0.97) |
| Female | 1380 | 139 | 1241 | 0.94 (0.91, 0.97) | 0.86 (0.80, 0.92) | 0.91 (0.89, 0.92) |
| Male | 667 | 90 | 577 | 0.94 (0.90, 0.97) | 0.83 (0.76, 0.91) | 0.93 (0.91, 0.96) |

**Supplementary Table 21.** Stratified holdout performance assessment for a literature-based COVID-19 susceptibility model based on two exposures and one symptom (HWF Exp + Symp)

| cohort | N | ncase | ncontrol | AUC | Sensitivity | Specificity |
| --- | --- | --- | --- | --- | --- | --- |
| Admixed African-European Genetic Ancestry | 96 | 12 | 84 | 0.94 (0.84, 1.00) | 0.92 (0.76, 1.00) | 0.94 (0.89, 0.99) |
| All | 2047 | 229 | 1818 | 0.90 (0.88, 0.93) | 0.78 (0.73, 0.83) | 0.94 (0.93, 0.95) |
| European Genetic Ancestry | 1457 | 146 | 1311 | 0.89 (0.86, 0.92) | 0.75 (0.68, 0.82) | 0.94 (0.93, 0.95) |
| Admixed Amerindian Genetic Ancestry | 155 | 23 | 132 | 0.94 (0.88, 1.00) | 0.87 (0.73, 1.00) | 0.92 (0.87, 0.97) |
| Other Genetic Ancestry | 339 | 48 | 291 | 0.92 (0.87, 0.97) | 0.79 (0.68, 0.90) | 0.96 (0.94, 0.98) |
| Ages 18-29 | 132 | 29 | 103 | 0.92 (0.85, 0.99) | 0.79 (0.65, 0.94) | 0.96 (0.92, 1.00) |
| Ages 30-49 | 582 | 77 | 505 | 0.91 (0.86, 0.95) | 0.82 (0.73, 0.91) | 0.92 (0.90, 0.95) |
| Ages 50-65 | 616 | 83 | 533 | 0.91 (0.87, 0.95) | 0.81 (0.73, 0.89) | 0.93 (0.91, 0.95) |
| Ages 65+ | 717 | 40 | 677 | 0.85 (0.77, 0.92) | 0.65 (0.50, 0.80) | 0.96 (0.95, 0.98) |
| Female | 1380 | 139 | 1241 | 0.91 (0.88, 0.94) | 0.81 (0.75, 0.88) | 0.94 (0.92, 0.95) |
| Male | 667 | 90 | 577 | 0.89 (0.85, 0.94) | 0.73 (0.64, 0.83) | 0.95 (0.94, 0.97) |

**Supplementary Table 22.** Stratified holdout performance assessment for a literature-based COVID-19 susceptibility model based on seven symptoms (HWF Symp)

| cohort | N | ncase | ncontrol | AUC | Sensitivity | Specificity |
| --- | --- | --- | --- | --- | --- | --- |
| Admixed African-European Genetic Ancestry | 96 | 12 | 84 | 0.90 (0.77, 1.00) | 0.84 (0.62, 1.00) | 0.92 (0.86, 0.98) |
| All | 2047 | 229 | 1818 | 0.87 (0.84, 0.90) | 0.75 (0.69, 0.80) | 0.91 (0.90, 0.93) |
| European Genetic Ancestry | 1457 | 146 | 1311 | 0.87 (0.83, 0.91) | 0.75 (0.68, 0.82) | 0.91 (0.89, 0.93) |
| Admixed Amerindian Genetic Ancestry | 155 | 23 | 132 | 0.92 (0.85, 0.99) | 0.87 (0.73, 1.00) | 0.93 (0.89, 0.97) |
| Other Genetic Ancestry | 339 | 48 | 291 | 0.83 (0.76, 0.91) | 0.67 (0.53, 0.80) | 0.92 (0.89, 0.95) |
| Ages 18-29 | 132 | 29 | 103 | 0.85 (0.75, 0.96) | 0.79 (0.64, 0.95) | 0.94 (0.90, 0.99) |
| Ages 30-49 | 582 | 77 | 505 | 0.89 (0.85, 0.93) | 0.75 (0.66, 0.85) | 0.88 (0.85, 0.91) |
| Ages 50-65 | 616 | 83 | 533 | 0.86 (0.80, 0.91) | 0.75 (0.65, 0.84) | 0.89 (0.87, 0.92) |
| Ages 65+ | 717 | 40 | 677 | 0.84 (0.76, 0.92) | 0.70 (0.55, 0.85) | 0.95 (0.93, 0.96) |
| Female | 1380 | 139 | 1241 | 0.87 (0.83, 0.91) | 0.76 (0.69, 0.83) | 0.90 (0.88, 0.92) |
| Male | 667 | 90 | 577 | 0.87 (0.83, 0.92) | 0.73 (0.63, 0.82) | 0.94 (0.92, 0.96) |

**Supplementary Table 23.** Comparison between train and holdout performances for different risk models.

| Model | Train AUC (95% CI) | Holdout AUC (95% CI) |
| --- | --- | --- |
| Susceptibility– no symptoms (Dem + Exp) | 0.83 (0.81, 0.85) | 0.84 (0.82, 0.84) |
| Susceptibility– with symptoms (Dem + Exp + Symp) | 0.95 (0.93, 0.97) | 0.94 (0.92, 0.96) |
| Susceptibility– literature-based "minimal" model (HWF Exp + Symp) | 0.90 (0.88, 0.92) | 0.90 (0.88, 0.93) |
| Susceptibility– literature-based "symptoms-only" model (HWF Symp) | 0.87 (0.84, 0.90) | 0.87 (0.84, 0.90) |
| Severity– hospitalization | 0.85 (0.82, 0.88) | 0.87 (0.84, 0.90) |
| Severity– critical case of COVID-19 | 0.90 (0.87, 0.93) | 0.90 (0.87, 0.93) |

Columns reflect mean and nominal 95% confidence intervals for the area under the curve (AUC) of trained models on either the training or holdout dataset. Confidence intervals were estimated via bootstrapping (Methods).

**Supplementary Table 24.** Stratified holdout performance assessment for models to predict hospitalization amongst COVID-19 positive cases.

| cohort | N | ncase | ncontrol | Auc | Sensitivity | Specificity |
| --- | --- | --- | --- | --- | --- | --- |
| Admixed African-European Genetic Ancestry | 110 | 20 | 90 | 0.81 (0.72, 0.90) | 0.85 (0.68, 1.00) | 0.71 (0.62, 0.80) |
| All | 1793 | 191 | 1602 | 0.87 (0.84, 0.90) | 0.83 (0.78, 0.88) | 0.75 (0.73, 0.77) |
| European Genetic Ancestry | 1133 | 120 | 1013 | 0.87 (0.84, 0.90) | 0.82 (0.76, 0.89) | 0.74 (0.72, 0.77) |
| Admixed Amerindian Genetic Ancestry | 219 | 16 | 203 | 0.94 (0.90, 0.98) | 0.93 (0.81, 1.00) | 0.79 (0.74, 0.85) |
| Other Genetic Ancestry | 331 | 35 | 296 | 0.85 (0.77, 0.93) | 0.80 (0.67, 0.94) | 0.75 (0.70, 0.80) |
| Ages 18-29 | 201 | 7 | 194 | 0.95 (0.91, 0.99) | 0.85 (0.57, 1.00) | 0.90 (0.85, 0.94) |
| Ages 30-49 | 705 | 49 | 656 | 0.87 (0.82, 0.93) | 0.76 (0.63, 0.88) | 0.85 (0.82, 0.88) |
| Ages 50-65 | 616 | 78 | 538 | 0.86 (0.81, 0.90) | 0.86 (0.78, 0.94) | 0.65 (0.61, 0.69) |
| Ages 65+ | 271 | 57 | 214 | 0.78 (0.72, 0.85) | 0.86 (0.77, 0.95) | 0.53 (0.47, 0.60) |
| Female | 1166 | 118 | 1048 | 0.87 (0.83, 0.90) | 0.80 (0.72, 0.87) | 0.77 (0.74, 0.79) |
| Male | 627 | 73 | 554 | 0.87 (0.83, 0.91) | 0.89 (0.82, 0.96) | 0.71 (0.67, 0.75) |

Case/control status is defined based on self-reported hospitalization due to COVID-19-related illness (Methods). Metric performance columns reflect mean and nominal 95% confidence intervals for each metric on an internal, independent holdout dataset (Methods). Cohort-level receiver operator characteristic (ROC) curves were constructed from predicted class probabilities of models trained on the entire survey cohort (Methods).

**Supplementary Table 25.**
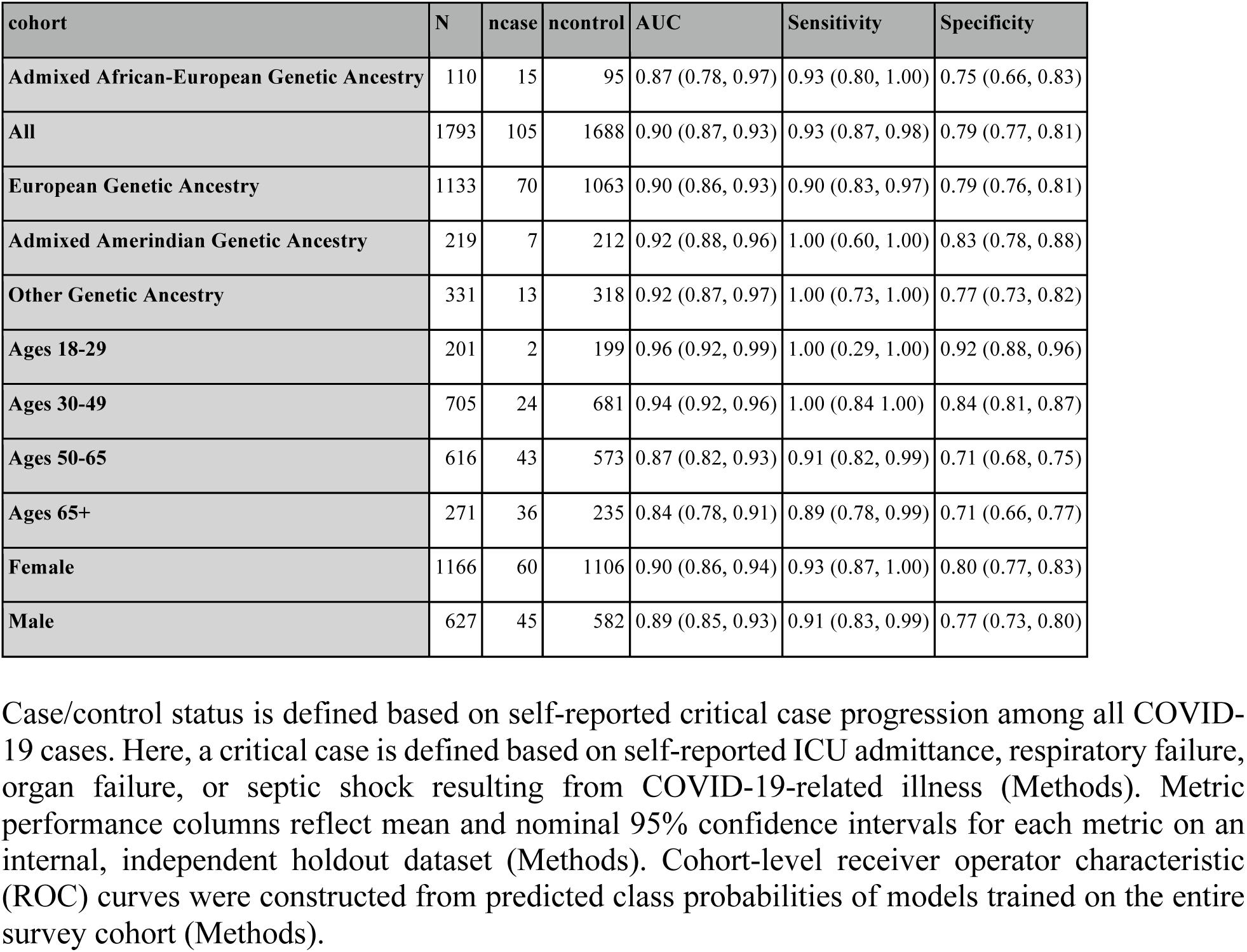
Stratified holdout performance assessment for models to predict critical COVID-19 cases amongst all COVID-19 positive cases.

| cohort | N | ncase | ncontrol | AUC | Sensitivity | Specificity |
| --- | --- | --- | --- | --- | --- | --- |
| Admixed African-European Genetic Ancestry | 110 | 15 | 95 | 0.87 (0.78, 0.97) | 0.93 (0.80, 1.00) | 0.75 (0.66, 0.83) |
| All | 1793 | 105 | 1688 | 0.90 (0.87, 0.93) | 0.93 (0.87, 0.98) | 0.79 (0.77, 0.81) |
| European Genetic Ancestry | 1133 | 70 | 1063 | 0.90 (0.86, 0.93) | 0.90 (0.83, 0.97) | 0.79 (0.76, 0.81) |
| Admixed Amerindian Genetic Ancestry | 219 | 7 | 212 | 0.92 (0.88, 0.96) | 1.00 (0.60, 1.00) | 0.83 (0.78, 0.88) |
| Other Genetic Ancestry | 331 | 13 | 318 | 0.92 (0.87, 0.97) | 1.00 (0.73, 1.00) | 0.77 (0.73, 0.82) |
| Ages 18-29 | 201 | 2 | 199 | 0.96 (0.92, 0.99) | 1.00 (0.29, 1.00) | 0.92 (0.88, 0.96) |
| Ages 30-49 | 705 | 24 | 681 | 0.94 (0.92, 0.96) | 1.00 (0.84, 1.00) | 0.84 (0.81, 0.87) |
| Ages 50-65 | 616 | 43 | 573 | 0.87 (0.82, 0.93) | 0.91 (0.82, 0.99) | 0.71 (0.68, 0.75) |
| Ages 65+ | 271 | 36 | 235 | 0.84 (0.78, 0.91) | 0.89 (0.78, 0.99) | 0.71 (0.66, 0.77) |
| Female | 1166 | 60 | 1106 | 0.90 (0.86, 0.94) | 0.93 (0.87, 1.00) | 0.80 (0.77, 0.83) |
| Male | 627 | 45 | 582 | 0.89 (0.85, 0.93) | 0.91 (0.83, 0.99) | 0.77 (0.73, 0.80) |

Case/control status is defined based on self-reported critical case progression among all COVID-19 cases. Here, a critical case is defined based on self-reported ICU admittance, respiratory failure, organ failure, or septic shock resulting from COVID-19-related illness (Methods). Metric performance columns reflect mean and nominal 95% confidence intervals for each metric on an internal, independent holdout dataset (Methods). Cohort-level receiver operator characteristic (ROC) curves were constructed from predicted class probabilities of models trained on the entire survey cohort (Methods).

### SUPPLEMENTARY FIGURES

**Supplementary Figure 1.**
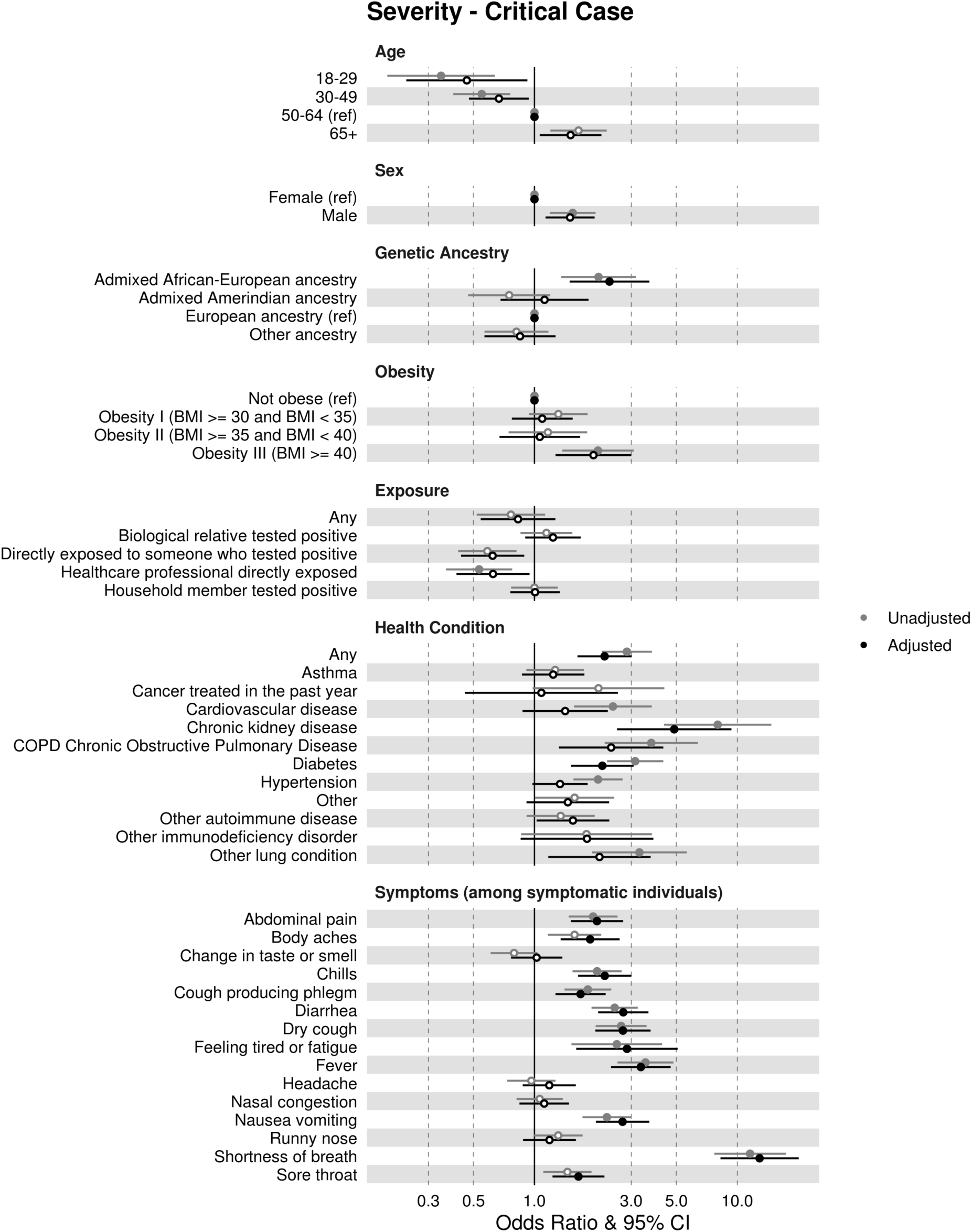
Severity (critical case) odds ratios (ORs) and 95% confidence intervals (CIs) estimated from simple (“Unadjusted models,” grey) and multiple (“Adjusted models,” black) logistic regression with adjustment for other risk factors. Open circles indicate not significant (p-value > 0.05) after accounting for multiple hypothesis tests using Bonferroni correction. Age, sex, genetic ancestry, and obesity ORs were estimated in relation to the reference variables indicated. Exposure, health, and symptom ORs were each estimated separately as binary variables. Symptom ORs were estimated as binary variables among symptomatic testers only (Methods). Risk factor adjustments for severity include: sex, age, obesity (Y/N), and underlying health conditions (Y/N if any). Where applicable, individual adjustment variables were omitted to avoid duplicate adjustment (Methods).

**Supplementary Figure 2.**
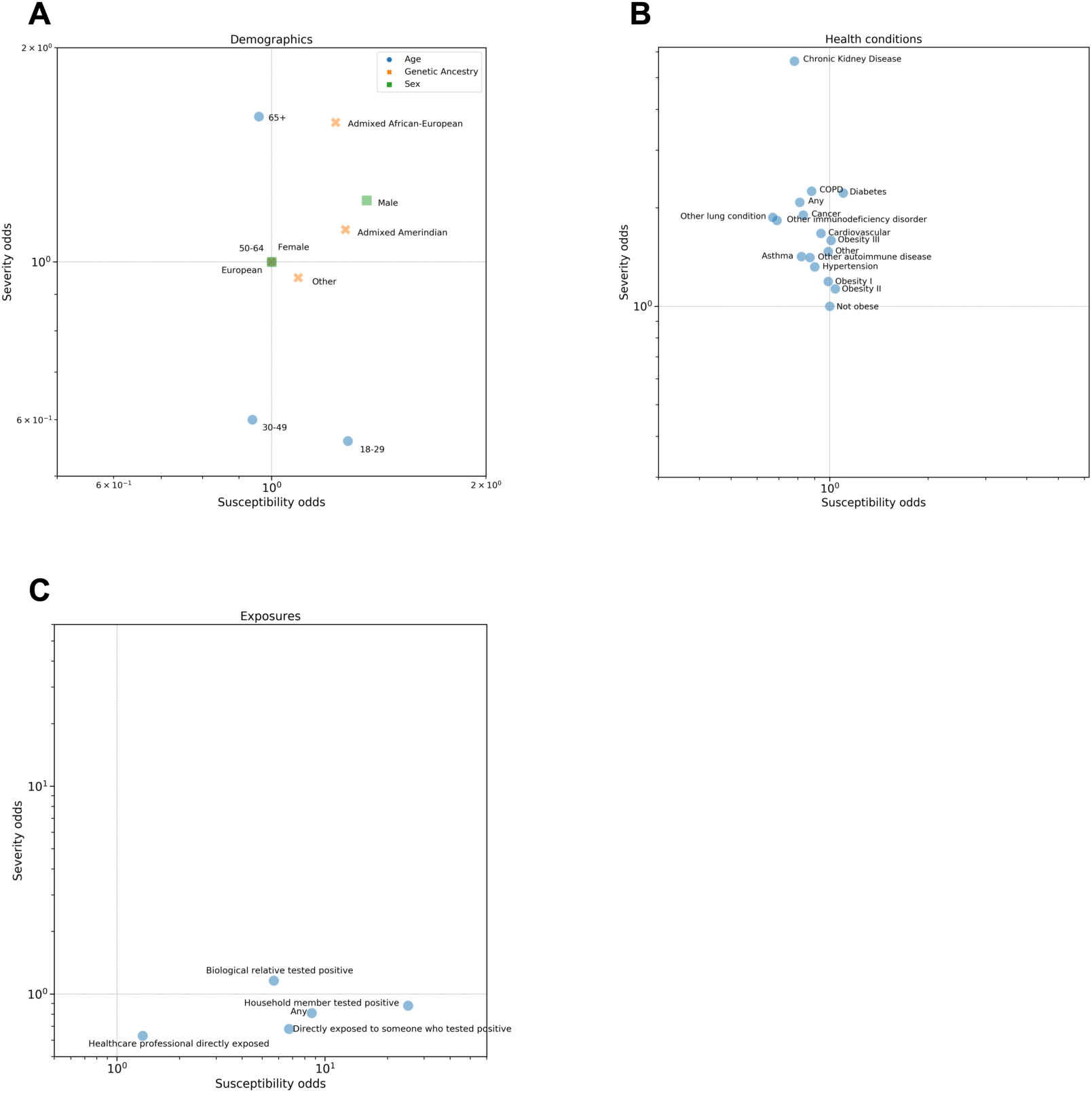
Comparison of Susceptibility and Severity adjusted ORs. Comparison of susceptibility ORs (horizontal axis) and severity ORs (vertical axis) from Figures 1 and 2. Severity aORs are for hospitalization. aORs are adjusted as in Figures 1 and 2 (see Figure 3 for adjusted ORs for symptoms). Plotted on a log scale for visibility. A) **Demographics**, broken down by age, sex, and genetic ancestry. B) **Health conditions**: Some pre-existing health conditions have increased odds of critical outcomes but susceptibility is not characterized by health conditions. C) **Exposures** are important for susceptibility but not severity.

**Supplementary Figure 3.**
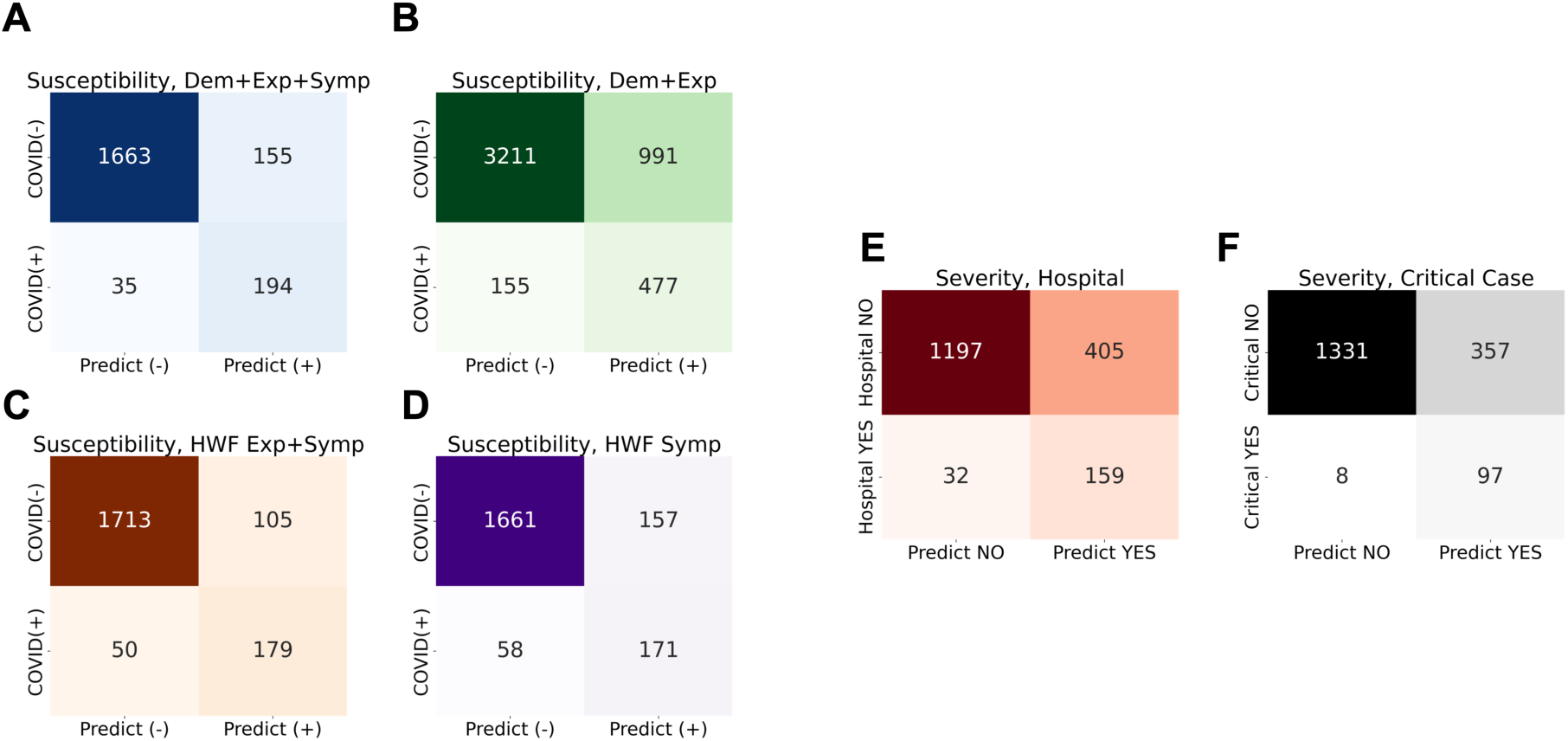
Risk model confusion matrices for independent holdout data. Numbers represent number of individuals in each category. A) Susceptibility model with demographics, exposures, and symptoms. B) Susceptibility model demographics and exposures (symptoms excluded). C) Literature-based susceptibility model with three risk factors including two exposures and change in taste or smell. D) Literature-based susceptibility model with seven COVID-19 symptoms. E) Hospitalization risk model. F) Critical case risk model. Refer to Methods as well as Supplementary Tables 18-25 for additional model performance data and model risk factor information.

**Supplementary Figure 4.**
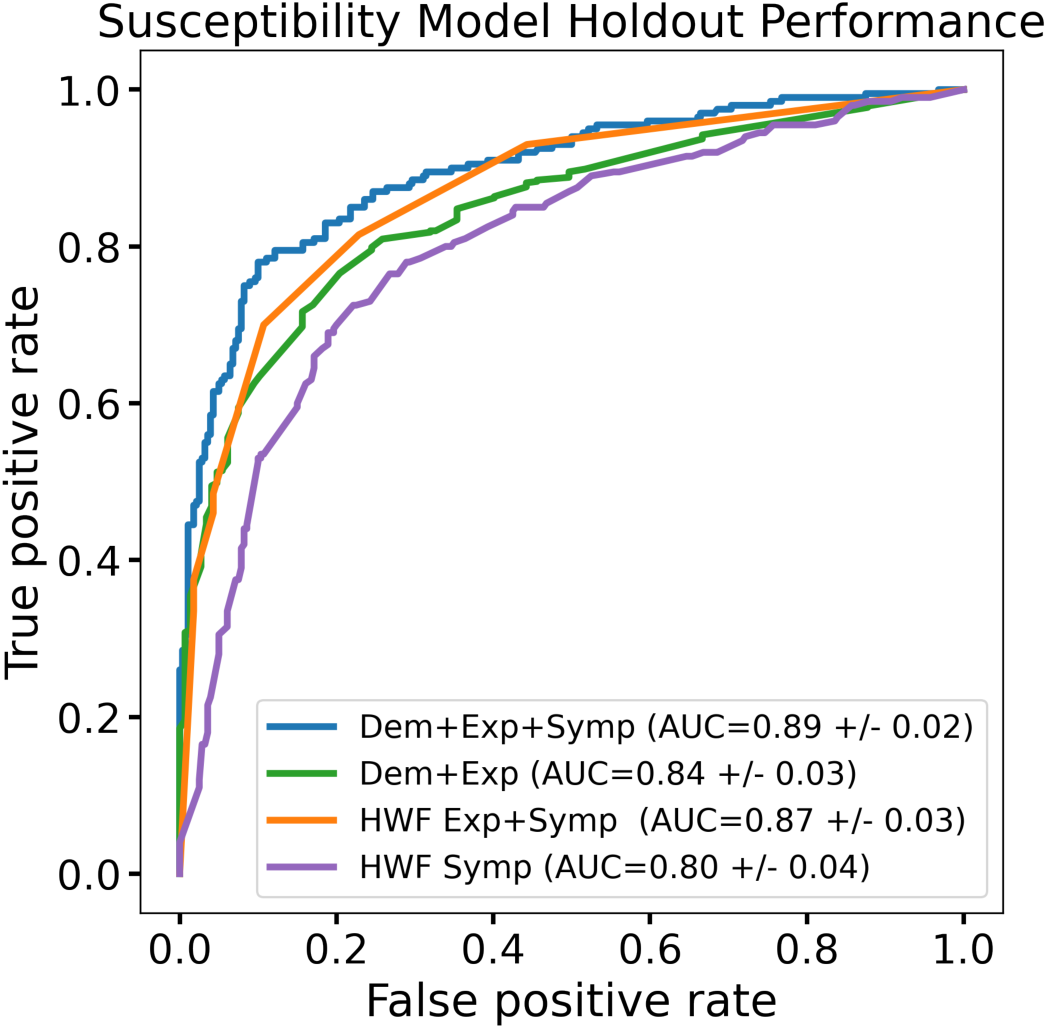
Performance of susceptibility models evaluated in a symptomatic cohort of positive and negative testers. Plot depicts receiver operating characteristic (ROC) curves for four susceptibility models (Supplementary Table 18). Here, the modeling cohort has been restricted to positive and negative testers reporting at least one symptom of moderate or greater intensity for model training and evaluation.

**Supplementary Figure 5.**
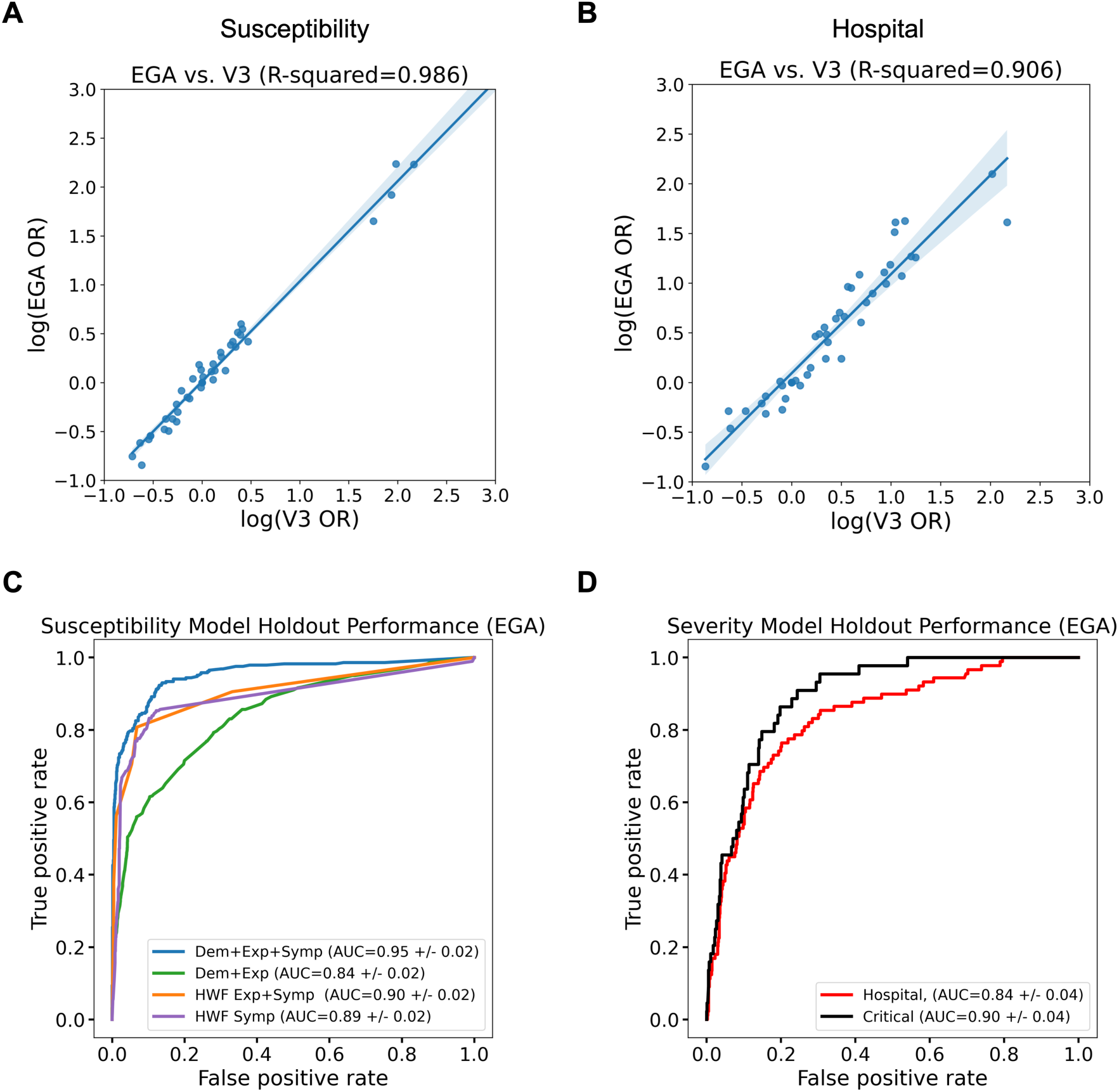
Concordance between the EGA cohort and AncestryDNA cohort (V3) from this study. A) Concordance of susceptibility associations between the EGA dataset (vertical axis) and the AncestryDNA dataset from this study (V3, horizontal axis), plotted as log mean effect estimates (logORs). B) Concordance of hospitalization associations between the EGA dataset (vertical axis) and the AncestryDNA dataset from this study (V3, horizontal axis). C) Independent holdout performance of susceptibility models trained within the EGA dataset. D) Independent holdout performance of severity models trained within the EGA dataset.

